# COVID-19 Myocarditis and Severity Factors: An Adult Cohort Study

**DOI:** 10.1101/2020.03.19.20034124

**Authors:** Kun-Long Ma, Zhi-Heng Liu, Chun-Feng Cao, Ming-Ke Liu, Juan Liao, Jing-Bo Zou, Ling-Xi Kong, Ke-Qiang Wan, Jun Zhang, Qun-Bo Wang, Wen-Guang Tian, Guang-Mei Qin, Lei Zhang, Fu-Jun Luan, Shi-Ling Li, Liang-Bo Hu, Qian-Lu Li, Hai-Qiang Wang

**Affiliations:** Central laboratory, Yongchuan Hospital of Chongqing Medical University, Xuanhua Road, No. 439, Yongchuan, Chongqing 402160, P.R. China; Department of Orthopedics, Yongchuan Hospital of Chongqing Medical University, Xuanhua Road, No. 439, Yongchuan, Chongqing 402160, P.R. China; Department of Orthopaedics, Xi’an Air Force 986 Hospital, Air Force Medical University, 172 Youyi Eastern Road, Xi’an 710054, P.R. China; Department of Radiology, Yongchuan Hospital of Chongqing Medical University, Xuanhua Road, No. 439, Yongchuan, Chongqing 402160, P.R. China; Yongchuan Center for Disease and Prevention, Huilong Avenue, No. 471, Yongchuan, Chongqing 402160, P.R. China; Department of Infectious disease, Yongchuan Hospital of Chongqing Medical University, Xuanhua Road, No. 439, Yongchuan, Chongqing 402160, P.R. China; Baoji Central Hospital, 8 Jiangtan Road, Baoji 721008, Shaanxi Province, P.R. China; School of Public Health, Xi’an Jiaotong University Health Science Center, Xi’an 710061, Shaanxi Province, P.R. China; Management Group of COVID-19, Yongchuan Hospital of Chongqing Medical University, Xuanhua Road, No. 439, Yongchuan, Chongqing 402160, P.R. China; Department of Respiratory Medicine, Yongchuan Hospital of Chongqing Medical University, Xuanhua Road, No. 439, Yongchuan, Chongqing 402160, P.R. China; Department of Medical Oncology, National Cancer Center/National Clinical Research Center for Cancer/Cancer Hospital, Chinese Academy of Medical Sciences & Peking Union Medical College, Beijing Key Laboratory of Clinical Study on Anticancer Molecular Targeted Drugs, Beijing 100021, China; Department of pediatrics, Yongchuan Hospital of Chongqing Medical University, Xuanhua Road, No. 439, Yongchuan, Chongqing 402160, P.R. China; Department of Neurology, Yongchuan Hospital of Chongqing Medical University, Xuanhua Road, No. 439, Yongchuan, Chongqing 402160, P.R. China; Institute of Integrative Medicine, Shaanxi University of Chinese Medicine, Xixian Avenue, Xixian District, Xi’an 712046, Shaanxi Province, P.R. China

## Abstract

**Background:** Notwithstanding the clinical hallmarks of COVID-19 patients were reported, several critical issues still remain mysterious, i.e., prognostic factors for COVID-19 including extrinsic factors as viral load of SARS-CoV-2 and intrinsic factors as individual’s health conditions; myocarditis incidence rate and hallmarks.

**Methods:** Demographic, epidemiologic, radiologic and laboratory data were collected by medical record reviews of adult hospitalized patients diagnosed as COVID-19. Cycle threshold (Ct) value data of real-time PCR (RT-PCR) were collected. The time duration was from 21 January to 2 March, 2020. Pulmonary inflammation index (PII) values were used for chest CT findings. Multivariate logistic regression analysis was used to identify independent severity risk factors.

**RESULTS:** In total, 84 hospitalized adult patients diagnosed as COVID-19 were included, including 20 severe and 64 nonsevere cases. The viral load of the severe group was significantly higher than that of the non-severe group, regardless of the Ct values for *N* or *ORF1ab* gene of virus (all p<0.05).Typical CT abnormalities was more likely existing in the severe group than in the nonsevere group in patchy shadows or ground glass opacities, consolidation, and interlobular septal thickening (all p<0.05). In addition, the PII values in the severe group was significantly higher than that in the nonsevere group (52.5 [42.5-62.5] vs 20 [5.0-31.6]; p<0.001). Amongst 84 patients, 13 patients (15.48%) were noted with abnormal electrocardiograms (ECGs) and serum myocardial enzyme levels; whereas 4 (4.8%) were clinically diagnosed as SARS-CoV-2 myocarditis. Multivariable logistic regress analysis distinguished three key independent risk factors for the severity of COVID-19, including age [OR 2.350; 95% CI (1.206 to 4.580); p=0.012], Ct value [OR 0.158; 95% CI (0.025 to 0.987); p=0.048] and PII [OR 1.912; 95% CI (1.187 to 3.079); p=0.008].

**Interpretation:** **T**hree key-independent risk factors of COVID-19 were identified, including age, PII, and Ct value. The Ct value is closely correlated with the severity of COVID-19, and may act as a predictor of clinical severity of COVID-19 in the early stage. SARS-CoV-2 myocarditis should be highlighted despite a relatively low incidence rate (4.8%). The oxygen pressure and blood oxygen saturation should not be neglected as closely linked with the altitude of epidemic regions.

**Research in context:** *Evidence before this study:* We searched Pubmed on March 15, 2020 using the terms (“COVID-19” OR “novel coronavirus” OR “2019 novel coronavirus” OR “2019-nCoV” OR “pneumonia” OR “coronavirus”), AND “Myocarditis” OR “Cycle threshold (Ct)” OR “Altitude”. We found that one article analyzed the risk factors affecting the prognosis of adult patients with COVID-19 in terms of survivorship, without considering Ct values as extrinsic factors. Moreover, there are no reported studies on viral myocarditis caused by COVID-19 and the relationship between the altitude and COVID-19.

*Added value of this study:* We retrospectively analyzed the clinical data, Ct values, laboratory indicators and imaging findings of 84 adult patients with confirmed COVID-19. Three key-independent risk factors of COVID-19 were identified in our study, including age [OR 2.350; 95% CI (1.206 to 4.580); p=0.012], Ct value [OR 0.158; 95% CI (0.025 to 0.987); p=0.048] and PII [OR 1.912; 95% CI (1.187 to 3.079); p=0.008]. Amongst 84 patients, 13 patients (15.48%) were noted with abnormal electrocardiograms (ECGs) and serum myocardial enzyme levels; whereas 4 (4.8%) were clinically diagnosed as SARS-CoV-2 myocarditis. Moreover, altitude should be considered for COVID-19 severity classification, given that oxygen partial pressure and blood oxygen saturation of regional patients vary with altitudes.

*Implications of all the available evidence:* **T**hree key-independent risk factors of COVID-19 were identified, including age, PII, and Ct value. The Ct value is closely correlated with the severity of COVID-19, and may act as a predictor of clinical severity of COVID-19 in the early stage. SARS-CoV-2 myocarditis should be highlighted despite a relatively low incidence rate (4.8%). The oxygen pressure and blood oxygen saturation should not be neglected as closely linked with the altitude of epidemic regions.

## Introduction

Since late 2019, a type of viral pneumonia named as COVID-19 has been greatly threatening people’s health and lives globally. Due to the epidemics of COVID-19 from Wuhan, Hubei Province to all parts of China and increasing number of countries,^1^ the medical community is facing with unprecedented challenges with swift actions all over the world. The pathogen of COVID-19 has been identified as SARS-CoV-2 (Severe acute respiratory syndrome coronavirus 2), a family member of coronaviruses.^2-4^

Wu and McGoogan^5^ reported the summary of 72, 314 COVID-19 cases in China from the point of Chinese Center for Disease Control and Prevention, including data until Feb 11, 2020. Notably, there were 416 cases aged less than 10 years (1%) and 549 cases aged between 10-19 years (1%). As early as Feb 7, 2020, Wang et al^6^ reported the clinical features of 138 patients in Wuhan, China, classifying cases into ICU-treated (26.1%) and ICU-free groups (73.9%). The median age was 56 years with 54.3% male patients. Patients treated in ICU were older, with underlying comorbidities, complaining of dyspnea and anorexia, in comparison with ICU-free patients. Laboratory indicators with statistical significance between two groups included higher levels of white blood cell and neutrophils counts, D-dimer, creatine kinase, and creatine. Notably, higher level of lactate dehydrogenase (LDH) was linked with ICU-treated group. Xu et al^7^ reported their clinical findings amongst 62 COVID-19 patients in Zhejiang Province, China. Due to limited number of patients admitted to ICU (1 case) and the less severity of cases, statistical significance of data was unavailable.

So far, Guan et al reported the clinical observational results with the largest number of COVID-19 patients as 1099 cases from 552 hospitals of 30 provinces of China.^8^ The majority of enrolled patients were over 65 years (41.9%) and nonsevere cases (926/1099). Moreover, Negative radiologic findings existed in 17.9% nonsevere patients and 2.9% severe patients. Various degrees of hemocytopenia presented on admission. Higher level of LDH was noted in severe group (58.1%) than nonsevere group (37.2%).

Hitherto, SARS-CoV-2 nucleic acid detection with real-time PCR (RT-PCR) is the diagnostic gold standard of COVID-19. Fortunately, the genome of SARS-CoV-2 was unraveled with 29903 nucleotides (MN908947.3)^3,4^, consisting of 11 genes. Taking advantage of suitable primers (recommended targeting *ORF1ab* gene from 266 to 21555 bp, and *N* gene from 28274 to 29533 bp, http://ivdc.chinacdc.cn/kyjz/202001/t20200121_211337.html), low copies of virus can be detected by PCR enriching through a number of PCR cycles. For RT-PCR diagnosis, the cycle threshold (Ct) value reflects viral copies, i.e., viral load. Lower Ct value represents higher viral load, as extrinsic factor for individuals. High viral load was reported in an asymptomatic 6-month-old infant with COVID-19, reflected by dynamic Ct value changes of RT-PCR for SARS-CoV-2.^5^ Despite high viral load reflected by low Ct values (13.73 for *ORF1ab* gene, 15.57 for *N* gene), the infant was asymptomatic. Notably, scarce evidence is available on viral load and clinical outcome, notwithstanding RT-PCR has been widely used in clinical practice.

Whereas these frontier studies presented critical lines of evidence on the emerging infectious disease as COVID-19, most patients were undergoing treatment with unclear outcome when data were summarized. Therefore, several critical issues still remain mysterious, i.e., prognostic factors for COVID-19 including extrinsic factors as viral load of SARS-CoV-2 and intrinsic factors as individual’s health conditions, including those identified as candidate risk factors (older age, underlying comorbidities, higher levels of white blood cell and neutrophils counts, D-dimer, creatine kinase, and creatine).^6^ Nevertheless, the clinical significance of abnormal serum myocardial enzymes identified by previous studies^6-8^ has not been well documented, in particular viral potentiality of myocardial damage and underlying mechanisms.

Accordingly, the study aimed for addressing these important issues based on a retrospective observational study of 84 adult cases in Chongqing municipality, China (adjacent to Hubei Province).

## Methods

### Data Sources

We performed a retrospective study highlighting the clinical and radiographic hallmarks of diagnosed cases as COVID-19 in Yongchuan, Chongqing from Jan 21 to Mar 2, 2020. As an officially nominated hospital for COVID-19 patients, Yongchuan Hospital (affiliated to Chongqing Medical University) has been in charge of the treatment of COVID-19 cases from Yongchuan District, Chongqing municipality (close to Hubei Province). The study was approved by the Ethics Review Committee of Yongchuan Hospital (No. 2020KLS-6). We obtained oral informed consent for involved patients and/or their relatives. All consecutive adult cases (aged over 18 years) diagnosed with COVID-19 were included. All patients were under close surveillance until Mar 2, 2020.

Demographic, epidemiologic, radiologic imaging and laboratory data were collected by intensive medical record reviews of included cases, with double checks and additional inquiries when appropriate. The diagnosis and treatment were conducted strictly conforming to the COVID-19 national program provided by the National Health Commission of China (5^th^ edition).^9^

### Pathogen confirmation with double SARS-CoV-2 tests

Each patient underwent double tests for SARS-CoV-2 nucleic acid detection every 24 hours on his/her nasopharyngeal swab samples. In brief, viral nucleic acid from samples was conducted using viral isolation kits (Daangene, Guangzhou, China). Fluorescence real-time PCR was performed with diagnostic reagent kits (two available kits; Sansure Biotech, Changsha, China and Daangene, Guangzhou, China) on LightCycler 480 Real-Time PCR instrument (Roche, Rotkreuz, Switzerland) at Yongchuan Disease Control Centre. Ct (Cycle threshold) value of no more than 40 (either genes or single gene with double tests) was considered as SARS-CoV-2 positive according to the manufacturer’s instructions. Thermal cycling was performed at 50 ° C for 15min, 95 °C for 15min, 45 cycles of 94 °C for 15s and 55 °C for 45s. The sensitivity of RT-PCR assay kits was 500copies/ml. The primers were: Primer 1 targeting open reading frame 1ab (*ORF1ab*) of SARS-CoV-2, 5’-CCCTGTGGGTTTTACACTTAA-3’; whereas primer 2 targeting nucleocapsidprotein(*N*) as 5’-GGGGAACTTCTCCTGCTAGAAT-3.^6-7^ Ct value data were collected from Yongchuan Disease Control Centre.

### Laboratory tests

Full spectrum laboratory tests were performed at admission and successive appropriate times, including blood count, serum biochemistry, serum inflammatory indicators, myocardial enzymes. Additional tests were performed according to patients’ detailed conditions of background diseases. Lymphocytes sorting analyses were conducted using flow cytometre (Bricyte e6, Mindray, Shenzhen, China) and pertaining assay kits (Mindray, Shenzhen, China).

### Chest CT scans

At admission, each patient underwent chest CT scans (Go.top, SIMENS; parameters, 120kv, 50-200mA, automatic modulation, thickness as 5mm) for the evaluation of lung lesions. Subsequently, follow-up chest CT scans were performed when suitable. Diagnostic images were analyzed by experienced radiologists. Pulmonary inflammation index (PII) was estimated according to the criteria proposed by Chongqing Radiologist Association of China,^10^ with distribution and size of lesions estimated. As for distribution, one lung segment equals one score with highest score as 20, representing left and right lung lesions. For lesion size, 1 score indicates over 50% lung segment volume invasive, whereas 0 score less than 50%. Accordingly, PII= (Distribution score+ Size score)/40*100%. PII values were calculated twice at three week intervals by two experienced radiologists. Interclass coefficient was used to assess inter-rater reliability.

### Electrocardiographic (ECG) examination

At admission, each patient underwent ECG examinations routinely. Each electrocardiogram was evaluated and reported by senior technicians. Abnormal ECGs were collected and analyzed in combination of clinical manifestations and serum myocardial enzyme tests. Repeated ECGs were ordered for patients suspected with myocardial damage.

### Statistical analysis

Data were recorded and summarized using spreadsheets. Patients in our cohort were classified into severe and nonsevere groups at admission in combination of national program and the American Thoracic Society guidelines.^11^ In brief, patients were included in severe group if they meet with the following criteria: 1) Shortness of breath with respiratory rate (RR) >30 times /min; 2) Oxygen saturation <93% at rest; 3)Arterial partial oxygen pressure (PaO2)/oxygen absorption concentration (FiO2)<277mmHg (The result was calculated according to the altitude of the epidemic area, Yongchuan city is about 700 meters above sea level, 1mmHg=0.133 kPa); 3) Pulmonary imaging showed that the lesion had progressed to > 50% at 24 ∼ 48 h; 4) Respiratory failure;5) Shock; 6)Combined with other organ failure. Continuous (means for normally distributed variables and/or interquartile ranges for abnormally distributed variables) and categorical variables (percentage, %) were analyzed using SPSS, version 29.0. Unpaired t test, Fisher exact test, Mann-Whitney U test and chi-square test were performed where appropriate. Multivariable logistic regress model was used to control for confounding factors and distinguish independent risk factors for clinical severity. Risk factors with a univariable p-value<0.05 were eligible for inclusion in the model. Severe group was coded as “1” and nonsevere group was coded as “0”. Variables (p≤0.10) were in the final model, with significant variables identified (p<0.05), via a forward and stepwise protocol. Adjusted odds ratios (ORs, indicating pertaining digital times more likely to present in severe group) and 95% confidence intervals (CIs) were calculated as independent risk factors. Spearman’s correlations were used to assess the links between multiple factors.

### Role of the funding source

No funding was received for the clinical cohort study. The corresponding author was in charge of all parts of the study and confirm that issues regarding the accuracy or integrity of the study had been properly scrutinized and settled. The final version of the manuscript was approved by all authors.

## Results

### Presenting Hallmarks

In total, the study cohort included 84 hospitalized adult patients diagnosed as COVID-19. By March 2, 2020, 68 of the 84 patients had been cured and discharged, and there were still 16 hospitalized patients with no deaths. The median age was 48 years (Interquartile Range [IQR], 42·3-62·5; range, 18-93) with 48 males (57.1%). Amongst these adult patients, 20 cases were classified as severe group at admission; whereas 64 cases as nonsevere group.

The median duration from illness onset to diagnosis was 2 days (IQR, 1-5). Onset symptoms were fever (54[64.3%]), cough (43[51.2%]), expectoration (28[33%]), fatigue (16[19.0%]), anorexia (14[16.7%]), myalgia (11[13.1%]), Dizziness(8[9.5%]), Pharyngalgia(8[9.5%]), Chilly(8[9.5%]), short of breath (7[8.3%]), diarrhea (6[7.1%]), Headache (4[4.8%]), Dyspnea(3[3.6%]), Nausea (3[3.6%]), Vomiting(2[2.4%]) and Fluster(2[2.4%]). Notably, there were 11(13.1%) asymptomatic cases in the cohort, and all were in the nonsevere group(table 1).

**Table 1.**
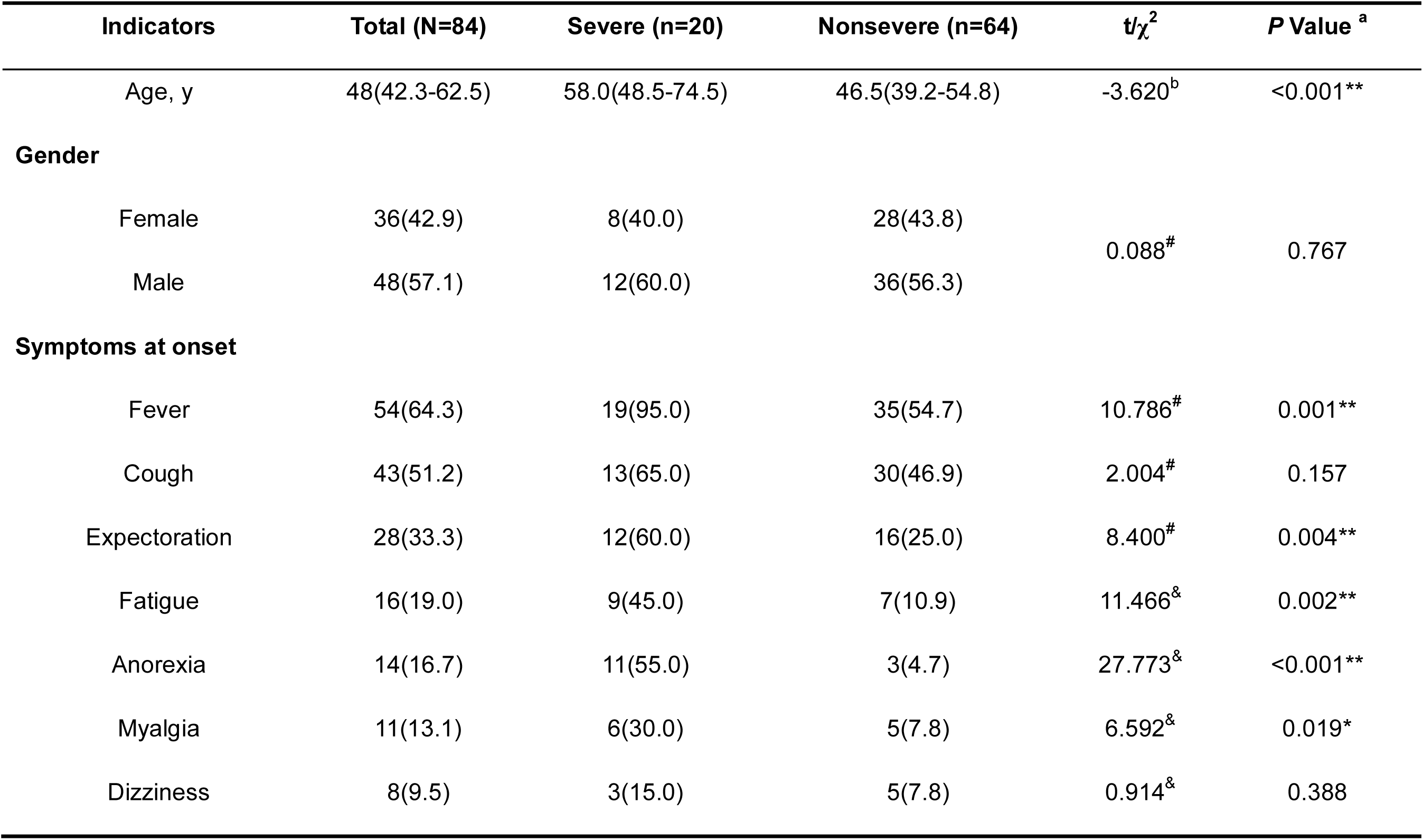
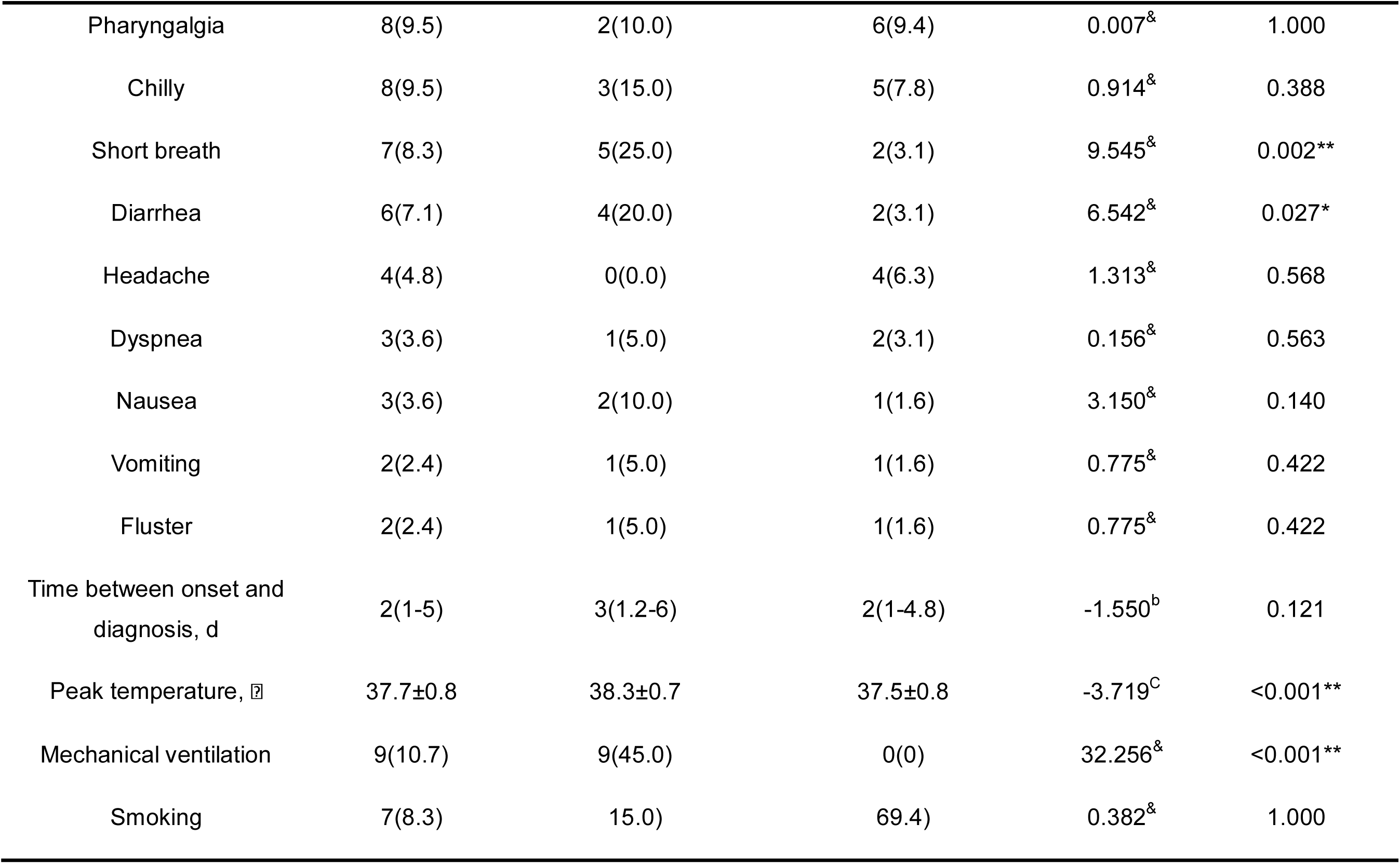
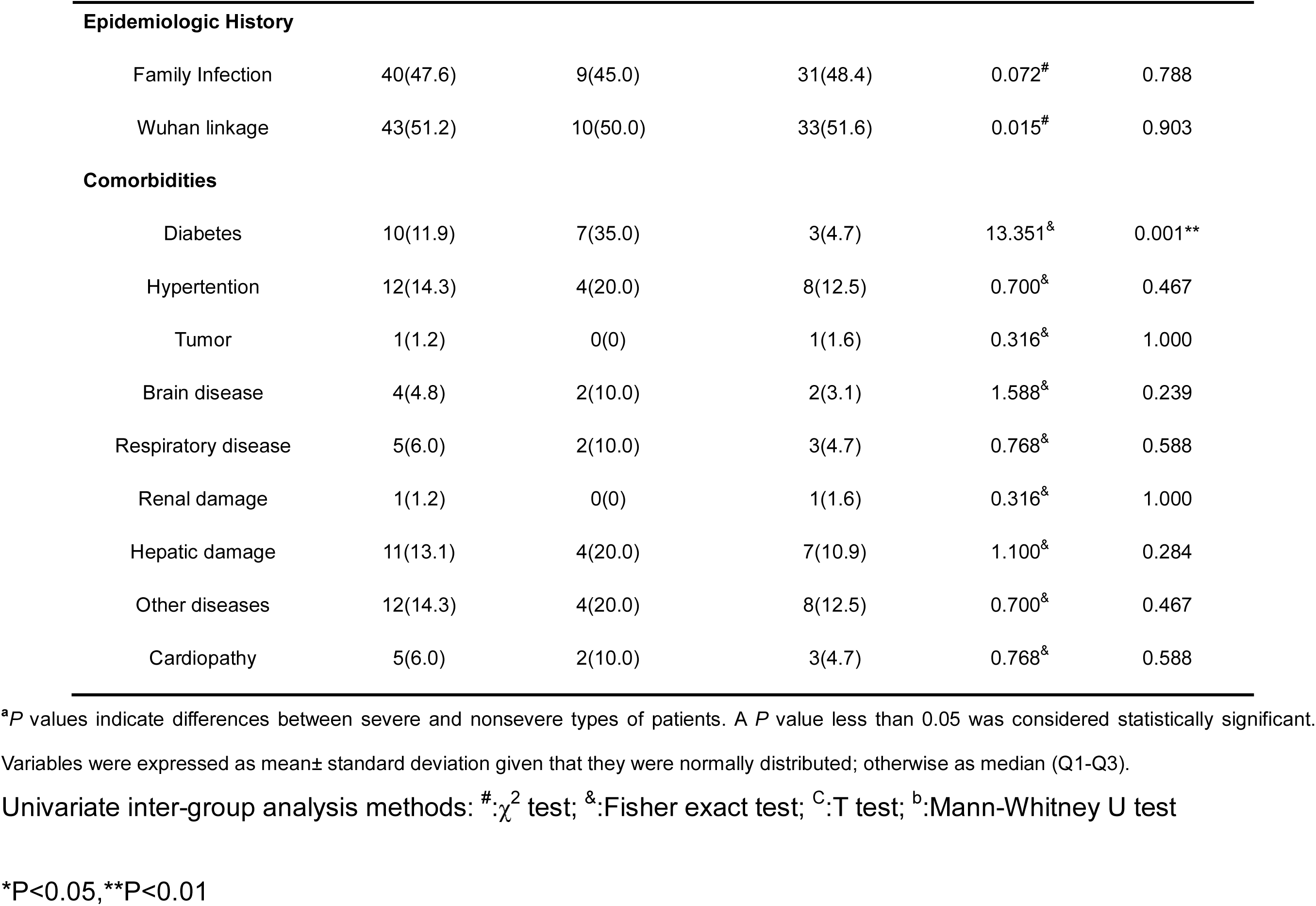
Baseline Hallmarks of Hospitalized Adult Patients With COVID-2019 (%)

The average peak temperature was 37.7°C. Forty cases (47.6%) had family cluster infection; whereas 43 patients (51.2%) had Wuhan linkage.

In comparison with nonsevere patients, the severe cases were older (58.0 [IQR, 48.5-74.5] vs 46.5 [IQR, 39.2-54.8], p<0.001), with higher peak temperatures ([38.3±0.7] vs [37.5±0.8]; p<0.001), and were more prone to symptoms such as fever (19[95.0%] vs 35[54.7%]; p<0.001), expectoration(12[60.0%] vs 16[25.0%]; p=0.002), fatigue (9[45.0%] vs 7[10.9%]; p=0.004), anorexia (11[55.0%] vs 3[4.7%]; p<0.001), myalgia (6[30.0%] vs 5[7.8%]; p=0.019), short breath (5[25.0%] vs 2[3.1%]; p=0.002) and diarrhea (4[20.0%] vs 2[3.1%]; p=0.027).

Thirty-five cases had one or more comorbidities. The most common coexisting diseases were hypertension (12[14.3%]), Hepatic damage (11[13.1%]), diabetes (10[11.9%]), cardiopathy (5[6.0%]), respiratory disease (5[6.0%]) and brain disease (4[4.8%]). Less common coexisting diseases were tumor (1[1.2%]) and renal damage (1[1.2%]). However, there was no significant difference in comorbidities except for diabetes (5[25%] vs 5[7.8%]; p=0.001).

### Laboratory indicators

There were significant differences in a number of laboratory indicators between patients in the severe and non-severe groups (table 2). Distinct inter-group laboratory findings included multitude measures listed(figure 1); (1)higher inflammatory markers, CRP (25.4 [IQR,15.4-35.5] vs 6.1[IQR, 1.6-12.6]; p<0.001), ESR (72.1[IQR,46.5-79.6] vs 31.0[IQR, 19.5-66.0]; p=0.002), PCT(0.07 [IQR,0.06-0.09] vs 0.05[IQR, 0.05-0.06]; p<0.001), and IL-6(13.9[IQR, 7.2-22.7] vs 2.8[IQR, 1.7-7.8]; p<0.001); (2)higher indicators of cardiac enzymes such as CK (77.3[IQR, 43.9-109.2] vs 51.1[IQR, 37.0-77.8]; p=0.015); LDH (529.1[IQR, 347.7-580.2] vs 385.5[IQR, 319.9-442.0]; p=0.004); α-HBDH (183.0[IQR, 154.7-224.8] vs 132.0[IQR, 118.4-154.3; p<0.001]), and IMA (75.3[IQR, 72.3-82.9] vs 69.2 [IQR, 65.2-73.6; p<0.001]); (3)higher levels of NEUT(5.0[IQR, 3.6-6.9] vs 3.9 [IQR, 2.9-4.8; p=0.004]), and lower levels of LYM (0.8[IQR, 0.6-1.2] vs 1.4[IQR, 1.1-1.7]; p <.001]); (4) other markers, SF(1104.0[IQR, 774.0-1370.5] vs 368.5 [IQR, 135.3-506.3; p<0.001]), PAB (175.6[IQR, 142.6-209.4] vs 227.5[IQR, 192.4-253.0; p=0.001]), GLB (7.0[IQR, 5.0-8.9] vs 5.7[IQR, 5.4-6.0; p=0.001]), and AST (27.7[IQR, 23.1-41.6] vs 21.8[IQR, 19.0-27.3; p=0.001]).

**Table 2.** Laboratory Findings of Hospitalized Adult Patients With COVID-2019.

| Indicators | Total (n=84) | Severe (n=20) | Nonsevere (n=64) | Z | P Value <sup>a</sup> |
| --- | --- | --- | --- | --- | --- |
| CRP (0-10mg/L) | 8.1(2.6-22.7) | 25.4(15.4-35.5) | 6.1(1.6-12.6) | -4.663 | <0.001** |
| ESR (0-20mm/h) | 42.3(23.3-74.5) | 72.1(46.5-79.6) | 31.0(19.5-66.0) | -3.115 | 0.002** |
| PCT (0-0.05ng/ml) | 0.06(0.05-0.07) | 0.07(0.06-0.09) | 0.05(0.05-0.06) | -4.740 | <0.001** |
| IL-6 (0-7pg/ml) | 4.6(1.8-11.8) | 13.9(7.2-22.7) | 2.8(1.7-7.8) | -3.910 | <0.001** |
| C3 (0.80-1.60g/L) | 1.3(1.2-1.5) | 1.4(1.3-1.5) | 1.3(1.2-1.4) | -1.661 | 0.097 |
| C4 (0.20-0.40g/L) | 0.31(0.26-0.39) | 0.32(0.30-0.43) | 0.31(0.26-0.39) | -1.109 | 0.268 |
| SAA (0-10mg/L) | 12.0(4.0-22.5) | 21.5(5.0-156.8) | 10.0(4.0-19.0) | -1.321 | 0.187 |
| D-dimer (0-0.5ug/ml) | 0.5(0.3-0.7) | 0.6(0.4-1.0) | 0.4(0.3-0.6) | -3.323 | 0.001** |
| PT (11.0-14.5S) | 12.8 ( 12.1-13.5) | 13.1(12.3-13.5) | 12.7(12.0-13.6) | -0.543 | 0.587 |
| CK (55-170U/L) | 53.5(39.1-82.3) | 77.3(43.9-109.2) | 51.1(37.0-77.8) | -2.437 | 0.015* |
| LDH (313-618U/L) | 395.5(325.0-466.5) | 529.1(347.7-580.2) | 385.5(319.9-442.0) | -2.920 | 0.004** |
| α-HBDH (72-182U/L) | 144.0(121.0-169.0) | 183.0(154.7-224.8) | 132.0(118.4-154.3) | -4.096 | <0.001** |
| CK-MB (0-25U/L) | 5.6(4.0-7.0) | 7.0(5.0-8.9) | 5.0(4.0-6.0) | -1.843 | 0.065 |
| cTnl (0-0.034ng/ml) | 0.016(0.012-0.021) | 0.016(0.012-0.020) | 0.017(0.012-0.021) | -0.743 | 0.457 |
| IMA (0-85U/ml) | 71.1(65.8-74.6) | 75.3(72.3-82.9) | 69.2(65.2-73.6) | -3.730 | <0.001** |
| SF (13-150ug/L) | 490.0(152.0-879.0) | 1104.0(774.0-1370.5) | 368.5(135.3-506.3) | -3.487 | <0.001** |
| PAB (200-400mg/L) | 218.5 ( 174.7-249.0 ) | 175.6(142.6-209.4) | 227.5(192.4-253.0) | -3.333 | 0.001** |
| ALT (9-50U/L) | 29.0(19.5-40.0) | 35.6(21.1-55.6) | 27.3(19.2-37.0 ) | -1.645 | 0.100 |
| AST (15-40U/L) | 23.0 ( 20.3-31.8 ) | 27.7(23.1-41.6) | 21.8(19.0-27.3) | -3.291 | 0.001** |
| BUN (2.86-8.20mmol/L) | 4.3(3.3-5.0) | 4.6(3.3-6.7) | 4.0(3.3-4.8) | -1.485 | 0.137 |
| CR (44-106mmol/L) | 64.7(53.3-76.6) | 62.4(50.3-77.0) | 64.7(54.3-76.5) | -0.532 | 0.594 |
| WBC (4-10x10<sup>9</sup>/L) | 6.0(5.1-6.9) | 6.4(5.4-8.5) | 5.8(4.9-6.8) | -1.644 | 0.100 |
| NEUT (1.8-6.3x10<sup>9</sup>/L) | 4.0(3.1-5.2) | 5.0(3.6-6.9) | 3.9(2.9-4.8) | -2.864 | 0.004** |
| LYM (1.1-3.2x10<sup>9</sup>/L) | 1.3(1.0-1.6) | 0.8(0.6-1.2) | 1.4(1.1-1.7) | -4.390 | <0.001** |
| LYM% (20-50%) | 23.4(18.3-28.4) | 14.4(8.4-22.5) | 25.0(21.2-30.0) | -4.621 | <0.001** |
| PLT (125-35010<sup>9</sup>/L) | 212.5(169.2-248.3) | 212.8(176.8-257.6) | 212.5(158.4-248.3) | -0.463 | 0.643 |
| GLB | 5.8(5.5-6.3) | 7.0(5.0-8.9) | 5.7(5.4-6.0) | -3.238 | 0.001** |
| Total T Lymphocyte(%) | 65.2(60.2-72.8 ) | 62.8(53.9-72.1) | 66.3(60.8-72.8) | -1.469 | 0.142 |
| Auxiliary /Inductive T lymphocyte (%) | 38.5(32.0-44.1 ) | 38.1(28.2-43.0) | 38.5(33.2-44.8) | -0.635 | 0.525 |
| Inhibitive/Cytotoxic T Lymphocyte (%) | 24.4(19.6-30.4 ) | 23.3(18.3-29.4) | 25.0(19.8-31.3) | -1.050 | 0.294 |
| Inhibitive/Cytotoxic T Lymphocyte count | 268.5(170.0-394.0 ) | 168.5(99.0-266.3) | 299.0(222.5-413.8) | -3.324 | 0.001** |
| Total T Lymphocyte count | 740.5(563.5-1073.8 ) | 540.0(278.8-746.8) | 832.0(631.5-1191.8) | -3.450 | 0.001** |
| Auxiliary /Inductive T lymphocyte count | 437.0(310.3-663.8 ) | 323.0(146.0-478.0) | 476.5(343.3-733.8) | -2.925 | 0.003** |
| Auxiliary/Inhibitive T Lymphocyte Ratio | 1.7(1.1-2.1 ) | 1.70(1.15-2.08) | 1.60(1.10-2.10) | -0.305 | 0.760 |
| B Lymphocyte (%) | 11.3(7.9-14.5 ) | 10.7(7.3-14.4) | 11.5(7.9-14.7) | -0.347 | 0.729 |
| NK (%) | 20.1(15.3-25.8 ) | 20.8(18.1-29.3) | 19.9(13.0-25.8) | -1.068 | 0.286 |
| NK count | 234.0(161.0-316.0 ) | 213.5(103.5-288.0) | 236.0(175.0-333.0) | -1.437 | 0.151 |
| B Lymphocyte count | 118.0(86.5-183.0 ) | 88.5(57.5-140.8) | 127.0(88.0-191.0) | -2.557 | 0.011* |

Abbreviations: CRP, C-reactive protein; ESR, erythrocyte sedimentation rate; PCT, procacitonin; IL-6, interleukin 6; C3 , complement 3; C4, complement 4; SAA, serum amyloid A; PT, prothrombin time; LDH, serum lactate dehydrogenase; CK, creatine kinase; α-HBDH, $\alpha$ -Hydroxybutyrate Dehydrogenase; CK-MB, creatine kinase isoenzymes; cTnI, cardiac troponin I; IMA, ischemia-modified albumin; BUN, blood urea nitrogen; CR, creatinine; WBC, white blood cell; NEUT, neutrophils; LYM , lymphocyte; PLT, blood platelet; PAB, prealbumin, SF, serum ferritin; GLB, glycosylated hemoglobin; NK, natural killer cell.
<sup>a</sup>*P* values indicate differences between severe and nonsevere types of patients. Variables were expressed as median (Q1-Q3).
Univariate inter-group analysis methods: #, $\chi^2$ test; &:Fisher exact test; <sup>b</sup>:Mann-Whitney U test;
\**P*<0.05, \*\* *P*<0.01

**Figure 1.**
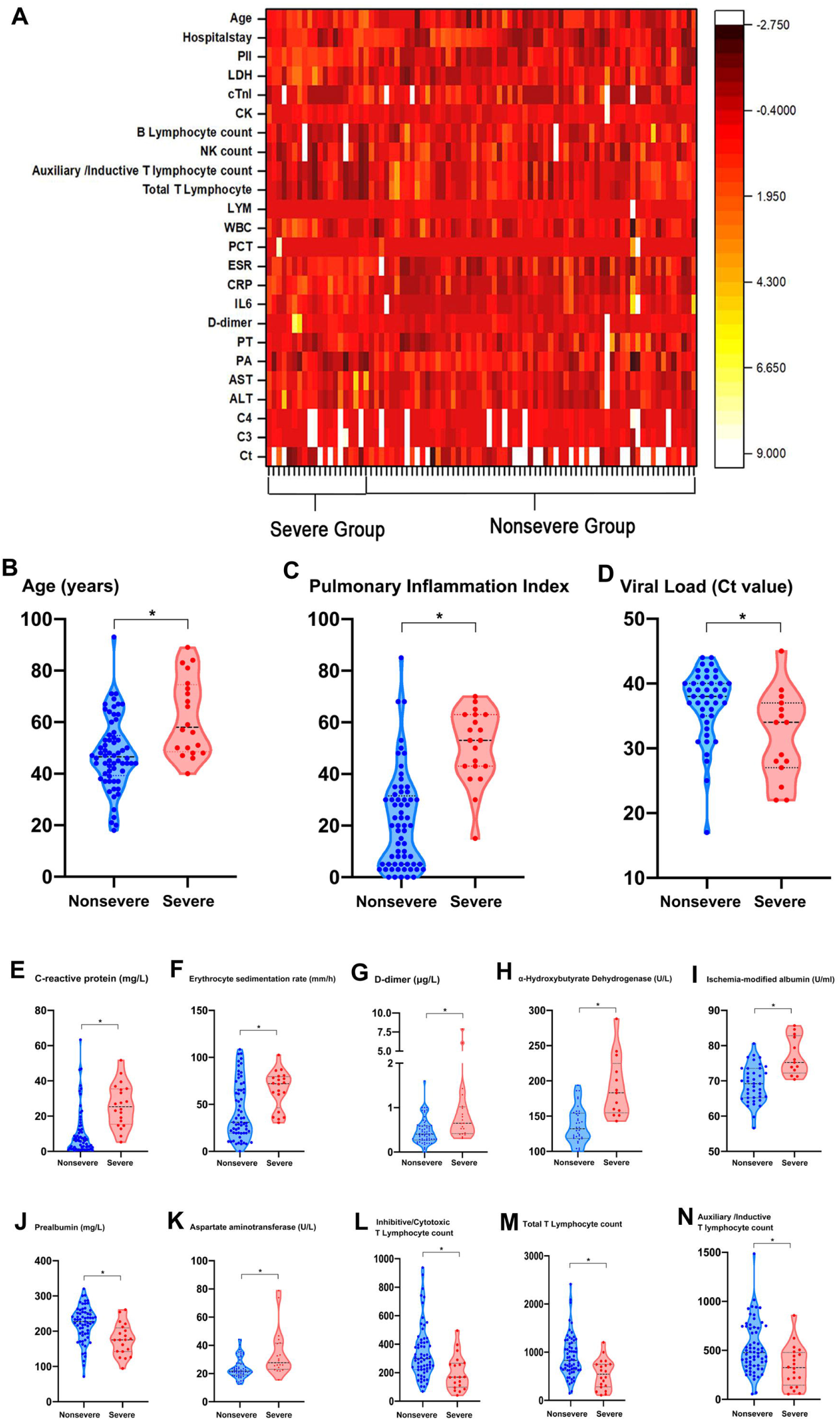
Distribution diagrams of risk factors and laboratory indicators with statistical significance. **A**, Heat map demonstrates the two-way hierarchical clustering of cases and Indictors. White strips without red or yellow colors indicate missing data. Yellow strips represent higher Z scores and red strip indicate lower Z scores. Severe and nonsevere groups represent the classification of COVID-19 according to clinical severity on admission. **B, C and D**, distribution diagrams of age, pulmonary inflammation index and viral load (Cycle threshold value) between severe and nonsevere groups, as identified independent risk factors for COVID-19 severity by multivariate logistic regression analysis. **E to N**, distribution diagrams of 10 laboratory indicators with statistical significance between severe and nonsevere groups by univariate analysis. \**p*<0.05.

In addition, in the detection of lymphocyte subsets, total T lymphocyte count, B lymphocyte count, and Inhibitive/Cytotoxic T Lymphocyte count were significantly reduced in the severe group compared with the nonsevere group.

### Ct values representing viral load

The Ct values of RT-PCR for two target genes (*ORF1ab* and *N*) were recorded in 62 of all patients, with a total of 207 values at corresponding time points (table 3). There were three different types of samples were used to detect SARS-CoV-2 infection, including 190 nasopharyngeal swab, 3 stool and 14 anal swab samples. In order to maintain the uniformity of the samples, the results of the anal swab samples and stool samples were excluded from our study. There were 14 patients with 51 nasopharyngeal swab samples in the severe group, while 48 patients with 139 nasopharyngeal swab samples in the nonsevere group. In the severe group, the median Ct values of RT-PCR for *N* gene was 33.8(IQR, 27.1-36.6) and for *ORF1ab* gene was 34.7(IQR, 30.9-37.7). While in the nonsevere group, the median Ct values of RT-PCR for N gene was 38.1(IQR, 34.2-40.2) and for *ORF1ab* gene was 38.6(IQR, 36.0-41.5). There was significant difference in Ct values either for *N* gene (p=0.004) or for *ORF1ab* gene (p=0.002) between the two groups.

**Table 3.**
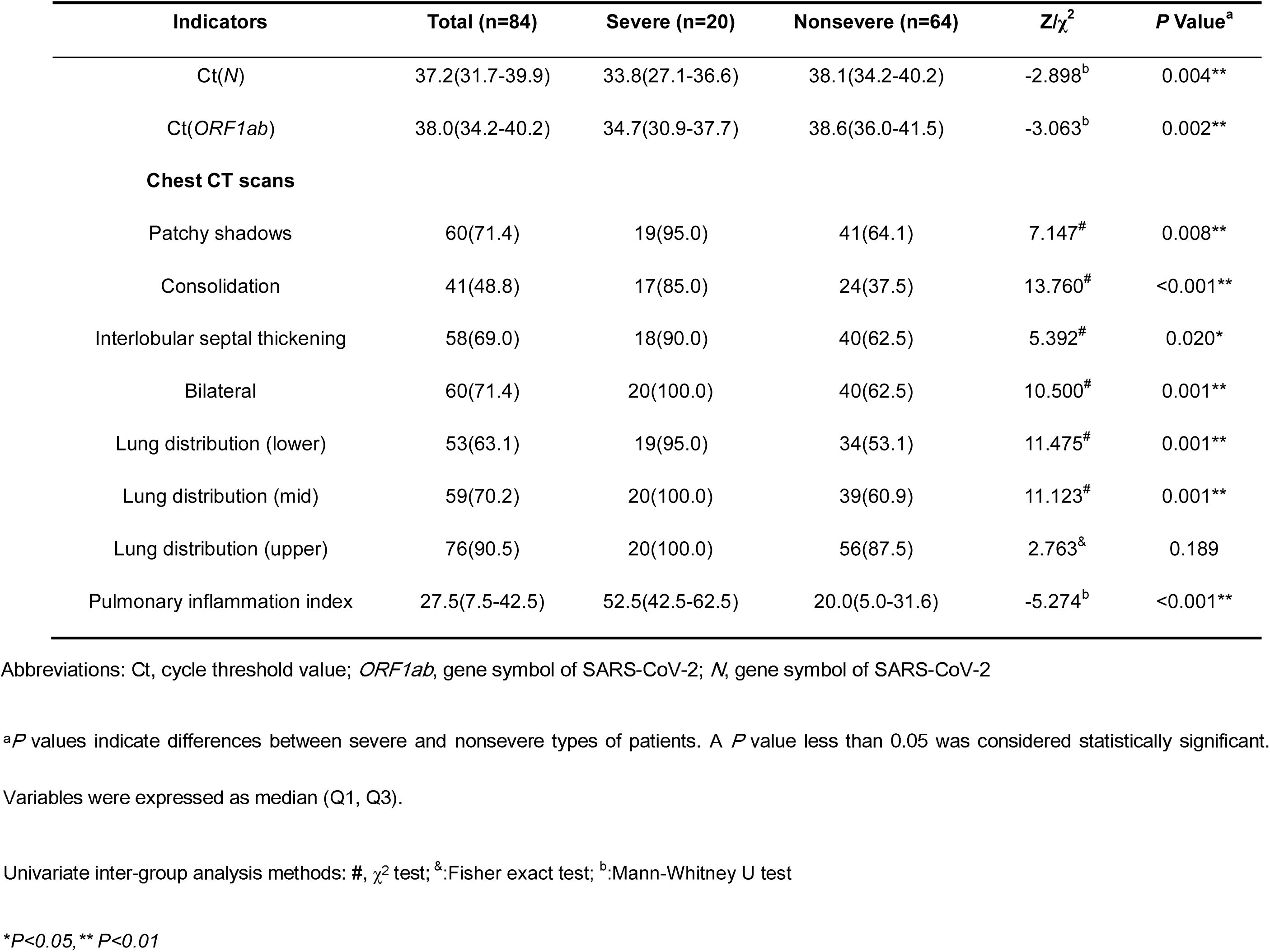
RT-PCR Ct Values and Chest CT findings of Hospitalized Adult Patients with COVID-19.

| Indicators | Total (n=84) | Severe (n=20) | Nonsevere (n=64) | Z/$\chi^2$ | P Value<sup>a</sup> |
| --- | --- | --- | --- | --- | --- |
| Ct(*N*) | 37.2(31.7-39.9) | 33.8(27.1-36.6) | 38.1(34.2-40.2) | -2.898<sup>b</sup> | 0.004** |
| Ct(*ORF1ab*) | 38.0(34.2-40.2) | 34.7(30.9-37.7) | 38.6(36.0-41.5) | -3.063<sup>b</sup> | 0.002** |
| **Chest CT scans** |  |  |  |  |  |
| Patchy shadows | 60(71.4) | 19(95.0) | 41(64.1) | 7.147<sup>#</sup> | 0.008** |
| Consolidation | 41(48.8) | 17(85.0) | 24(37.5) | 13.760<sup>#</sup> | <0.001** |
| Interlobular septal thickening | 58(69.0) | 18(90.0) | 40(62.5) | 5.392<sup>#</sup> | 0.020* |
| Bilateral | 60(71.4) | 20(100.0) | 40(62.5) | 10.500<sup>#</sup> | 0.001** |
| Lung distribution (lower) | 53(63.1) | 19(95.0) | 34(53.1) | 11.475<sup>#</sup> | 0.001** |
| Lung distribution (mid) | 59(70.2) | 20(100.0) | 39(60.9) | 11.123<sup>#</sup> | 0.001** |
| Lung distribution (upper) | 76(90.5) | 20(100.0) | 56(87.5) | 2.763<sup>&</sup> | 0.189 |
| Pulmonary inflammation index | 27.5(7.5-42.5) | 52.5(42.5-62.5) | 20.0(5.0-31.6) | -5.274<sup>b</sup> | <0.001** |

Abbreviations: Ct, cycle threshold value; *ORF1ab*, gene symbol of SARS-CoV-2; *N*, gene symbol of SARS-CoV-2
<sup>a</sup>*P* values indicate differences between severe and nonsevere types of patients. A *P* value less than 0.05 was considered statistically significant.
Variables were expressed as median (Q1, Q3).
Univariate inter-group analysis methods: #, $\chi^2$ test; &; Fisher exact test; <sup>b</sup>:Mann-Whitney U test
\**P*<0.05, \*\* *P*<0.01

### Chest CT findings and PII

Totally 84 patients diagnosed with COVID-19 underwent at least two or more CT examinations (table 3). An excellent inter-rater reliability was noted in the assessment of PII values(>0.89).The median of pulmonary inflammation index (PII) was 27.5(IQR, 7.5-42.5). The study showed that the PII values in the severe group was significantly higher than that in the nonsevere group (52.5 [42.5-62.5] vs 20 [5.0-31.6]; p<0.001). The most common CT abnormalities were patchy shadows or ground glass opacities (60[71.4%]), consolidation (41[48.8%]), interlobular septal thickening (58 [69%])(figure 2). Most patients have multiple areas of lung lesions with bilateral lung lesions (60[71.4%]), lower lung distribution (53[63.1%]), mid lung distribution (59 [70.2%]), and upper lung distribution (76[90.5%])(figure 3). Typical CT abnormalities was more likely existing in the severe group than in the nonsevere group with significant difference in patchy shadows or ground glass opacities (19[95%] vs 41[64.1%]; p=0.008), consolidation (17[85%] vs 24[37.5]; p<0.001), and interlobular septal thickening (18[90.0%] vs 40[62.5%]; p=0.020).There were also significant differences in the areas of lung lesions between the two groups. Compared with the nonsevere group, patients in the severe group were more likely to have bilateral lung lesions (20[100%] vs 40[62.5%]; p=0.001), and the lesions was more frequent in the lower lobe (19[95 %] vs 34[53.1%]; p=0.001) and middle lobe (20 [100 %] vs 39[60.9%]; p=0.001) of the lung(figure 4). However, no significant difference was found in the upper lobe of the lung (20[100 %] vs 56 [87.5%]; p=0.189).

**Figure 2.**
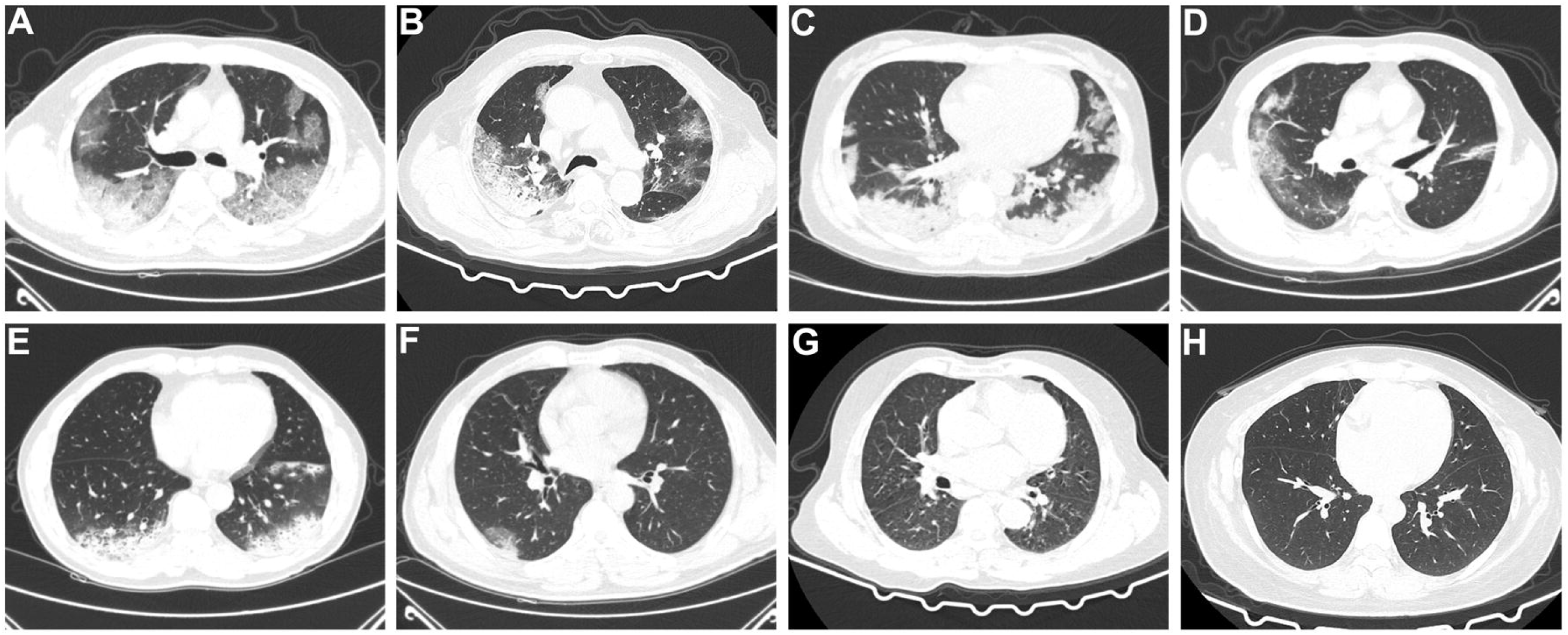
Representative chest CT images of COVID-19 patients. **A, B, C** and **D**, severe type. **E, F, G** and **H**, nonsevere type. **A**, a middle-aged male patient, chest CT scan performed on 12th days of onset. Multiple diffused ground glass opacities, dense shadow cord strip and nodules in both lung fields were revealed. **B**, an elderly male patient, chest CT scan performed on 11th days of onset. CT showed bilateral lung multiple diffused ground glass opacities and consolidation, with local interlobular septal thickening and crazy paving appearance. **C**, a middle-aged female patient, chest CT scan performed on 7th days of onset. CT showed bilateral subpleural multiple diffused ground glass opacities and consolidation. **D**, a middle-aged male patient, chest CT scan performed on 5th days of onset, multiple diffused ground glass opacities, dense shadow cord strip and nodules in bilateral subpleural lung regions, with bronchiectasis in the subpleural region of the lateral basal segment of the right lower lobe. **E**, chest CT scan performed on 18th days of onset, a male patient aged 54. Patchy lung infiltrates showed in bilateral subpleural lung regions, with intralesional low-density lesion. **F**, a middle-aged male patient, chest CT scan performed on 4th days of onset, A few fibrotic foci in the upper lobe of the right lung after clinical cue of TB. **G**, chest CT scan performed on 6th days of onset, an elderly male patient. CT showed a few fibrotic foci in the right middle lobe medial segment. **H**, chest CT scan performed on 5th days of onset, a young female patient. CT showed a few fibrotic foci in the medial segment of right middle lobe of lung.

**Figure 3.**
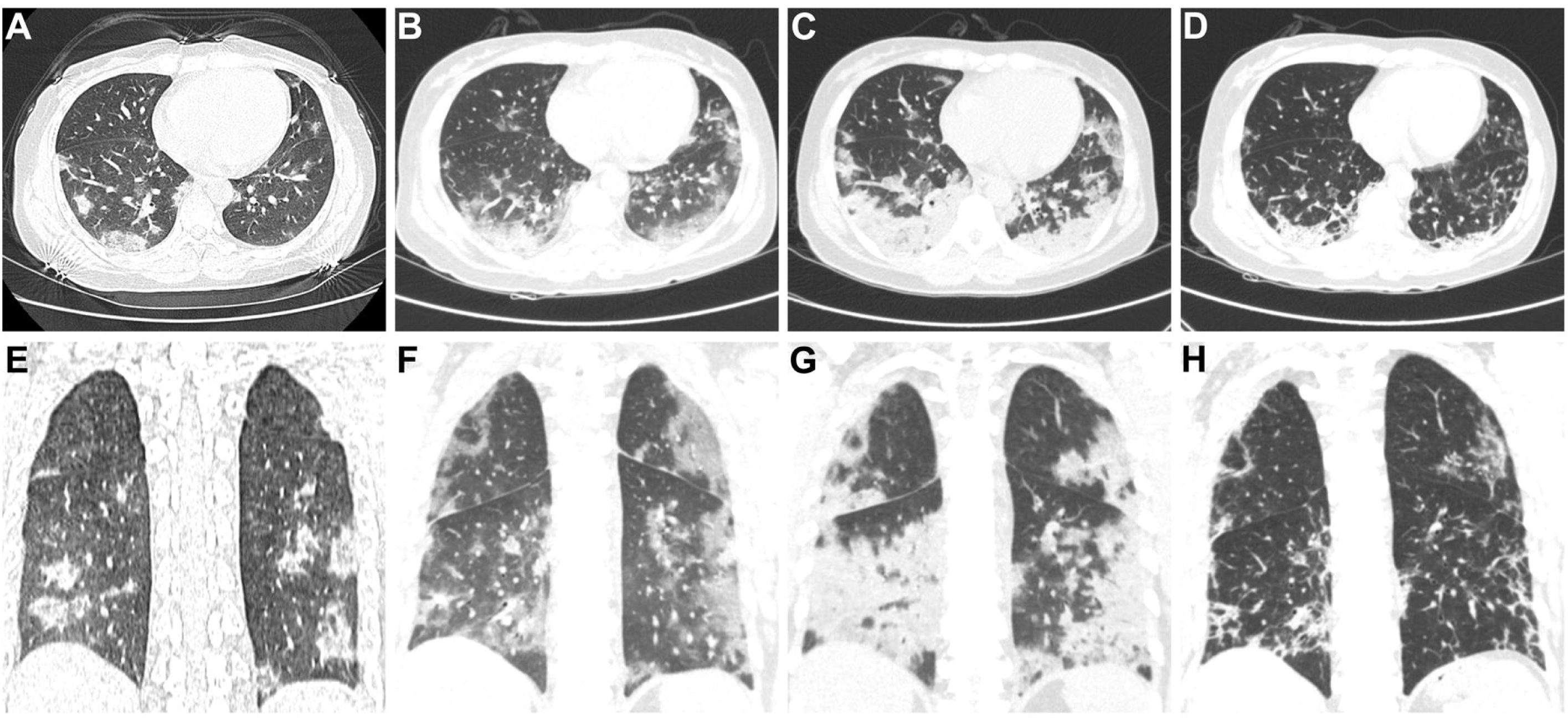
Dynamic chest CT scan changes of a nonsevere patient (a young male patient) with COVID-19. **A** and **E**, early-stage of COVID-19, chest CT scan performed on 2th days of onset. Irregular nodular high-density shadow in the basal segment of the left lower lobe. **B** and **F**, lung progressions, chest CT scan performed on 5th days of onset. CT showed enlarged lesion area, with patchy slightly high-density shadow and intralesional interlobular septal thickening. **C** and **G**, critical period, chest CT scan performed on 9th days of onset. CT showed enlarged lesion area from subpleural area to the hilum of lung, with lung consolidation and bronchiectasis. **D** and **H** convalescence phase, chest CT scan performed on 14th days of onset. CT showed decrease of the *density focus and* consolidation area, with gross fibrotic foci emerging.

**Figure 4.**
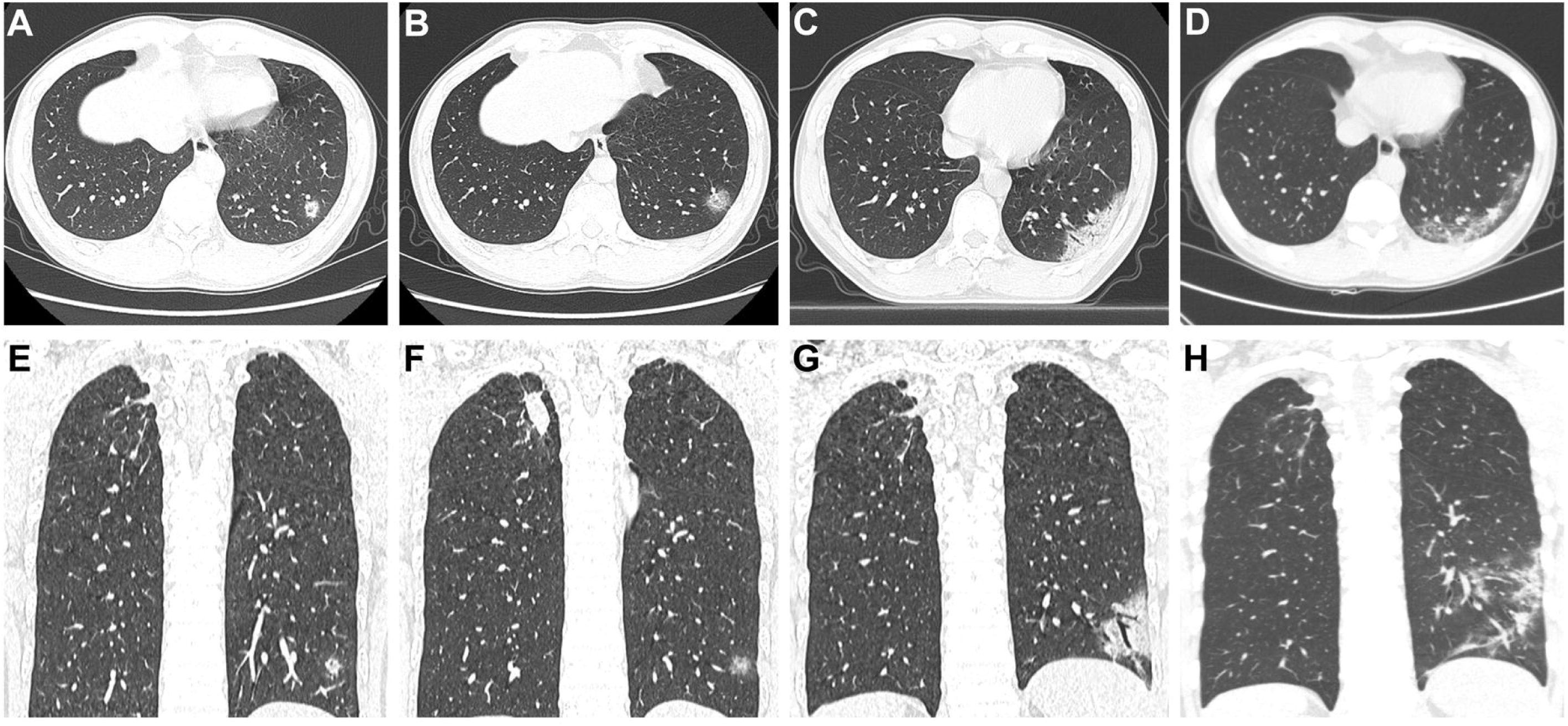
Dynamic chest CT scan changes of a severe patient (a middle-aged female patient) with COVID-19. **A** and **E**, early-stage of COVID-19, chest CT scan performed on 3th days of onset. CT showed ground-glass opacities and patchy lung infiltrates in bilateral subpleural lung regions with intralesional interlobular septal thickening. **B** and **F**, lung progressions, chest CT scan performed on 7th days of onset. CT showed enlarged lesion area, with patchy slightly high density shadow. **C** and **G**, critical period, chest CT scan performed on 11th days of onset. CT showed slightly enlarged lesion area and increase of lesion density. **D** and **H**, convalescence phase, chest CT scan performed on 20th days of onset. CT showed reduction of lesion and decrease of lesion density, with lesion absorption and fibrotic foci emerging.

### Complications during hospitalization

Complications during hospitalization were documented (table 4). There were 47 (56.0%) cases with electrolyte disorders, 37(44.0%) cases with anemia, 36(42.9%)cases with abnormal cardiac enzymes, 30(35.7%)cases with hepatic abnormalities, 27(32.1%)cases with hypoproteinemia, 23(27.4%) cases with gastrointestinal disorders, 22(26.2%) cases with bacterial pneumonia, 20(23.8%) cases with type 1 respiratory failure, and 6(7.1%) cases with renal abnormalities. Except for gastrointestinal disorders, the incidence of all complications in the severe group was significantly higher than that in the non-severe group (p<0.05). Of the 20 patients with respiratory failure, 9(45%) were treated with assisted mechanical ventilation, 7(35%) with nasal catheters and 4(20%) with masks. The median time from onset to respiratory failure was 7.75 days and to mechanical ventilation was 8 days.

**Table 4.** Complications during hospitalization of Adult Patients With COVID-2019(%)

| Complications | Total (N=84) | Severe (n=20) | Nonsevere (n=64) | $\chi^2$ | P Value <sup>a</sup> |
| --- | --- | --- | --- | --- | --- |
| Anemia | 37(44.0) | 13(65.0) | 24(37.5) | 4.676<sup>#</sup> | 0.031* |
| Hypoalbuminemia | 27(32.1) | 16(80.0) | 11(17.2) | 27.564<sup>#</sup> | <0.001** |
| Hepatic abnormality | 30(35.7) | 13(65.0) | 17(26.6) | 9.806<sup>#</sup> | 0.002** |
| Renal abnormality | 6(7.1) | 5(25.0) | 1(1.6) | 12.620<sup>&</sup> | 0.003** |
| Cardiac enzyme abnormality | 36(42.9) | 15(75.0) | 21(35.6) | 11.074<sup>#</sup> | 0.001** |
| Electrolyte disturbance | 47(56.0) | 19(95.0) | 28(43.8) | 16.240<sup>#</sup> | <0.001** |
| Respiratory failure | 20(23.8) | 20(100.0) | 0(0) | 13.440<sup>&</sup> | 0.003** |
| Intestinal dysbacteriosis | 23(27.4) | 6(30.0) | 17(26.6) | 0.091<sup>#</sup> | 0.763 |
| Bacterial pneumonia | 22(26.2) | 10(50.0) | 12(18.8) | 7.698<sup>#</sup> | 0.006** |
| Viral myocarditis | 4(4.8) | 3(15.0) | 1(1.6) | 6.067<sup>&</sup> | 0.040* |

<sup>a</sup>P values indicate differences between severe and nonsevere types of patients. A P value less than 0.05 was considered statistically significant.
Univariate inter-group analysis methods: #, $\chi^2$ test; &, Fisher exact test;
*\* $P < 0.05$ , \*\* $P < 0.01$*

### Clinical diagnosed myocarditis

Among all 84 patients, we found 13 patients with abnormal myocardial injury indicators combined with abnormal ECG, including 8 severe patients and 5 nonsevere patients (table 5). Four of the 13 patients had a history of cardiovascular diseases, while the other 9 patients had no such history. There were 3 cases with a history of hypertension, 1 patient with hypertension, atrial fibrillation and coronary heart disease who was excluded from our further analysis. The 12 remaining patients had abnormal changes in the myocardial enzymes or cTnI, amongst which 10 cases with high levels of inflammatory markers (except for patient no.9 and 11), and eleven of them exhibiting clinical symptoms of myocardial injury (except for patient no. 9).

**Table 5.**
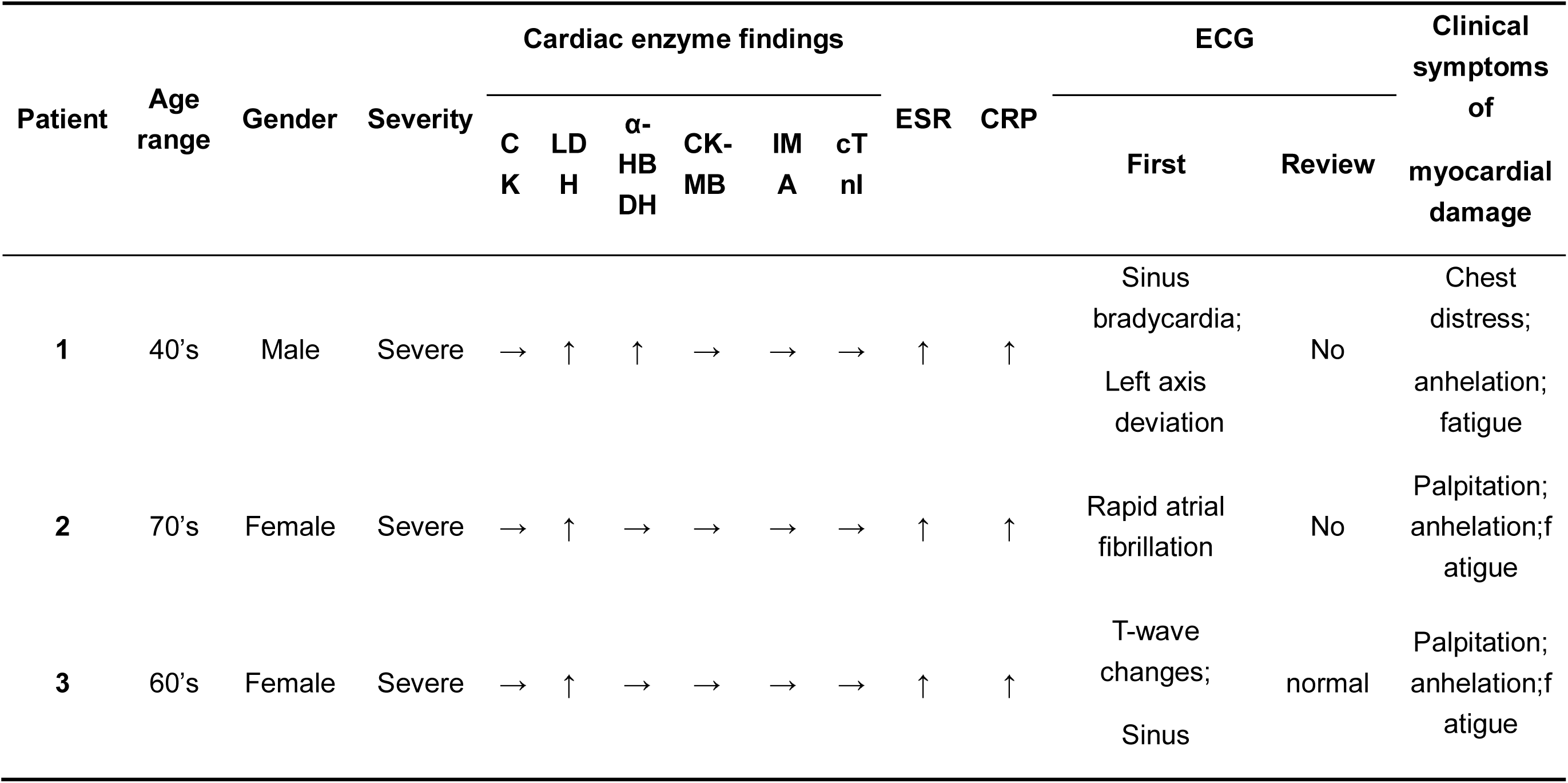
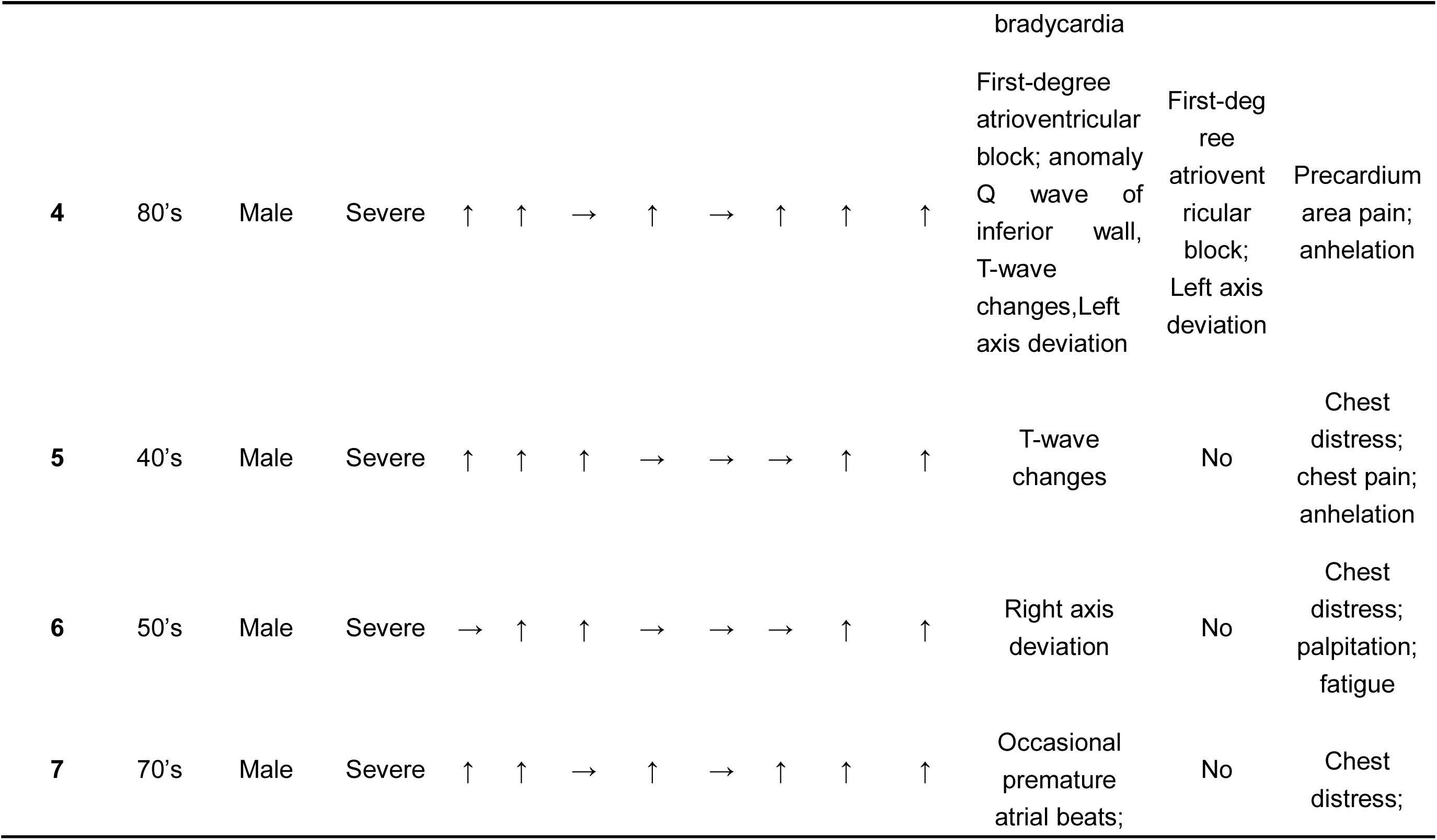
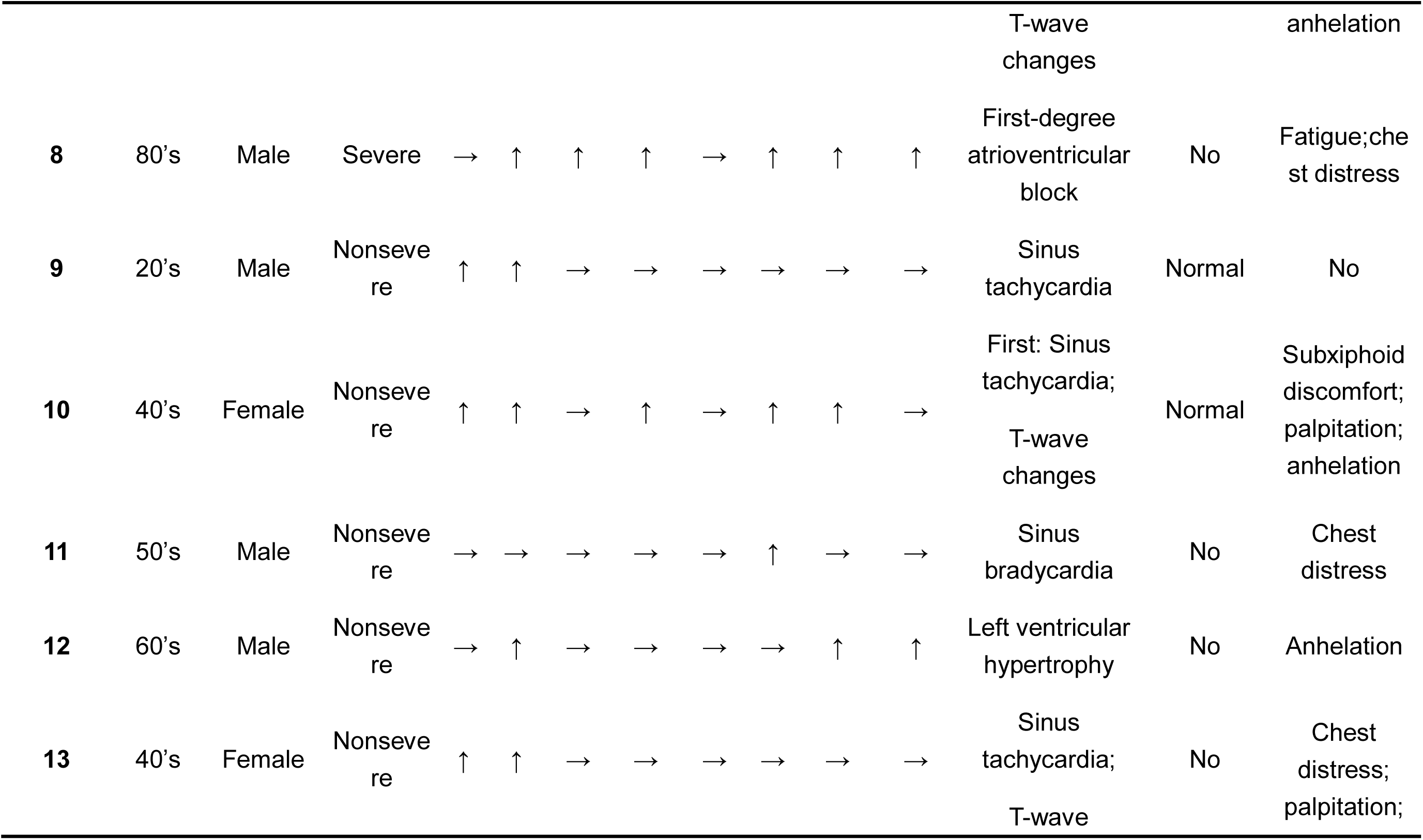
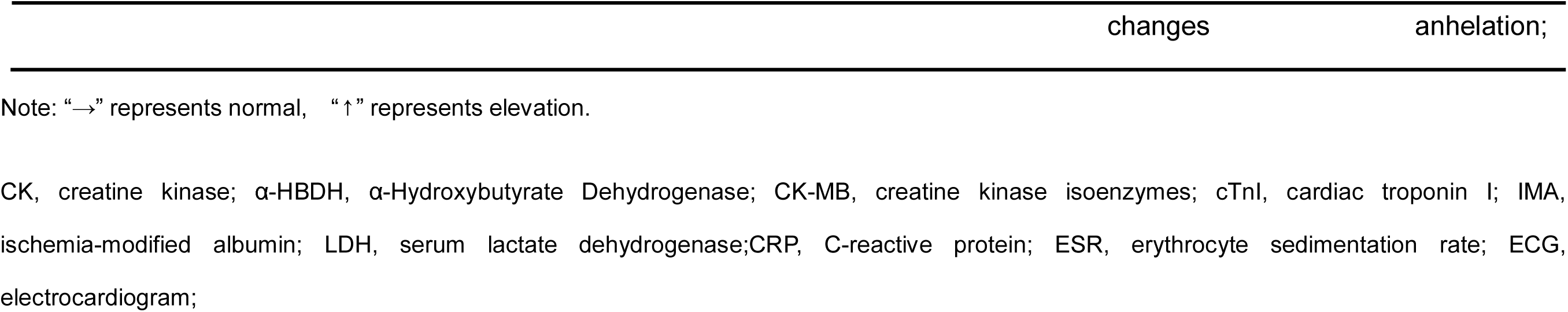
Cardiac Laboratory Findings for Patients with Abnormal ECG.

Typical abnormal ECG findings were noted in ten patients (except for patient no.2, 6 and 12). Major ECG abnormalities included sinus tachycardia or bradycardia, t-wave changes, atrioventricular block, and anomaly Q wave. Four patients underwent repeated ECG examinations. Patient no. 3 was an elderly female patient in the severe group. The first ECG indicated t-wave changes and sinus bradycardia. However, repeated ECG restored normal after treatment with a decrease in inflammatory markers and cardiac enzyme levels. Patient no. 4 was an elderly male patient in the severe group. His first ECG indicated First-degree atrioventricular block, t-wave changes, left axis deviation and abnormal Q wave in the lower wall, while the abnormal Q and t-wave disappeared after treatments. Patient no. 9 and 10 were in the nonsevere group. Their initial ECGs showed sinus tachycardia and sinus tachycardia associated with t-wave changes respectively, with repeated normal ECGs after treatment.

Based on the most updated diagnostic criteria of viral myocarditis^12-13^ (Disease code: H00395), 4 cases(patient no.4, 7, 8 and 10) were clinically diagnosed as SARS-CoV-2 myocarditis in combination of their manifestations, i.e., confirmed SARS-CoV-2 infection, typical ECG abnormalities, and elevated laboratory myocardial damage indicators(figure 5)

**Figure 5.**
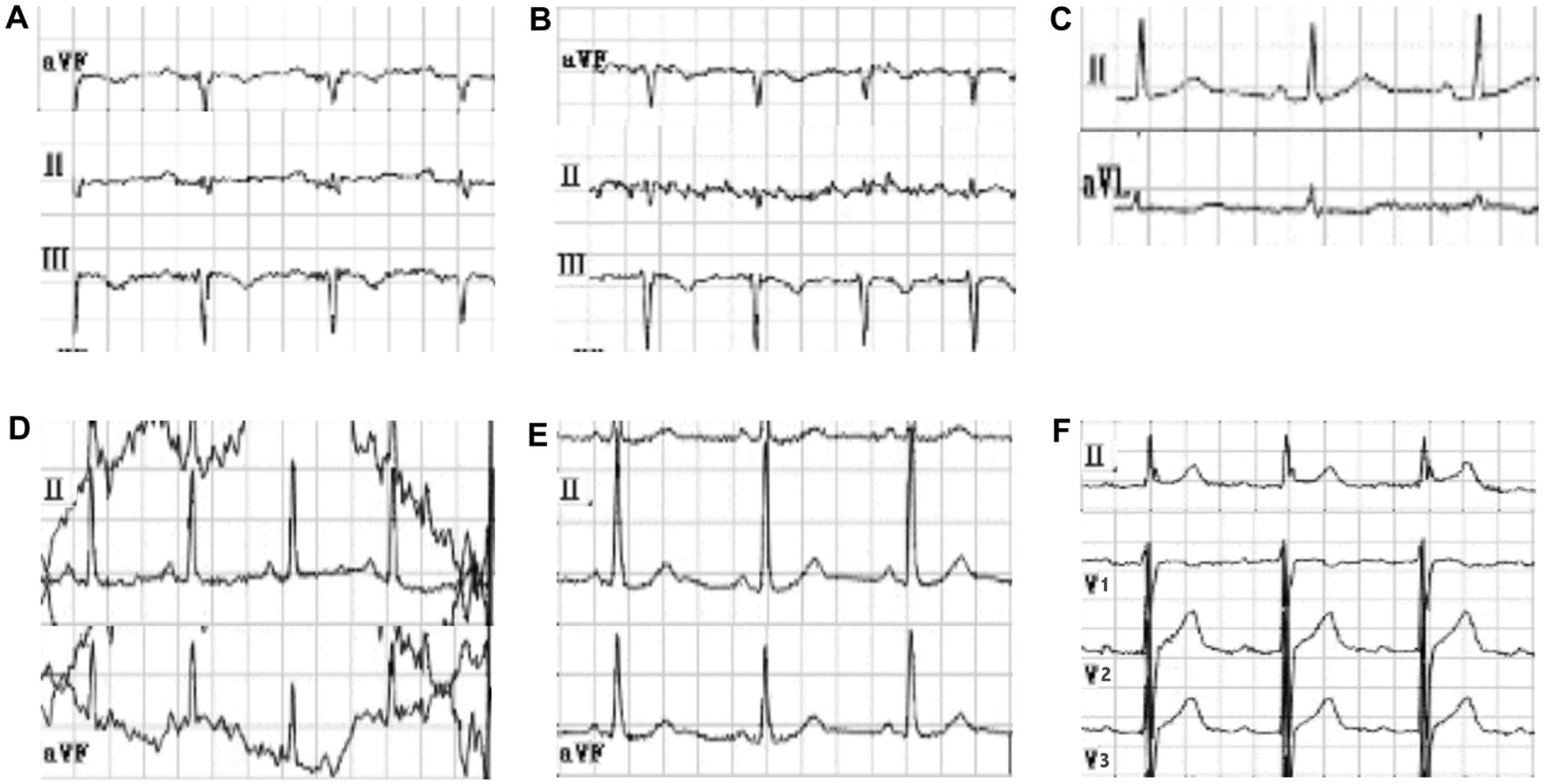
Representative electrocardiograms of clinical diagnosed SARS-CoV-2 myocarditis. **A**, Abnormal ECG of patient no. 4 on the day of admission. It showed low and flat t-wave in II lead, first-degree atrioventricular block and abnormal Q wave in the II, III, and aVF leads. **B**, ECG of patient no. 4 after treatments. It showed the abnormal Q and t-wave disappeared after treatments, leaving him with only an first-degree atrioventricular block in II, III, and aVF leads. **C**, Abnormal ECG of patient no. 7 on the day after admission. It showed occasional premature atrial beats in II lead, and inverted t-wave in aVL lead. **D**, Abnormal ECG of patient no. 10 after admission. It showed sinus tachycardia associated with low and flat t-wave in II lead and sinus tachycardia associated with positive and negative t-wave in aVF lead. **E**, ECG of patient no. 10 patient returned to normal in II and aVF leads after treatment. **F**, Abnormal ECG of patient no. 8 after admission. It showed first-degree atrioventricular block in II, V1, V2, and V3 leads.

In order to explore the intrinsic correlations between cardiac enzymes, ECG and cTn I with inflammatory cytokines and lymphocyte levels, we used Mann-Whitney U test to analyze the relevant data (table 6).The results showed that the level of cardiac enzymes was significantly positively correlated with the level of CRP (p=0.004) and PCT (p=0.012). In addition, this study also found that there was a significant correlation between the abnormality of ECG and the level of PCT (p=0.004) and lymphocytes counts (p=0.041). However, we did not find any correlation between cTn I and levels of inflammatory cytokines or lymphocytes.

**Table 6.** Cardiac Enzymes and ECG of Hospitalized Adult Patients with COVID-19.

| Indicators | Total | Abnormal | Normal | Z | P value |
| --- | --- | --- | --- | --- | --- |
| **Cardiac enzymes** | 84 | 36 | 48 |  |  |
| CRP | 8.08(2.63-22.73) | 14.70(5.42-35.49) | 6.68(2.08-14.67) | -2.852 | 0.004** |
| IL-6 | 4.59(1.82-11.80) | 7.43(2.37-18.36) | 3.26(1.75-8.39) | -1.926 | 0.054 |
| ESR | 42.25(23.25-74.50) | 62.00(30.28-77.75) | 32.25(19.75-66.00) | -1.576 | 0.115 |
| PCT | 0.055(0.050-0.070) | 0.058(0.050-0.080) | 0.050(0.050-0.067) | -2.517 | 0.012** |
| LYM counts | 1.32 ( 1.00-1.61 ) | 1.26 ( 0.79-1.56 ) | 1.33 ( 1.07-1.63 ) | -1.451 | 0.147 |
| Total T Lymphocyte counts | 740.50(563.50-1073.75) | 681.50(466.00-988.75) | 803.50(639.50-1174.75) | -1.921 | 0.055 |
| **cTn I** | 84 | 9 | 75 |  |  |
| CRP | 8.08(2.63-22.73) | 2.80(1.20-36.02) | 8.22(2.93-2.03) | -0.651 | 0.515 |
| IL-6 | 4.59(1.82-11.80) | 2.40(1.50-10.30) | 4.97(2.01-12.92) | -1.361 | 0.173 |
| ESR | 42.25(23.25-74.50) | 62.67(19.13-70.67) | 42.00(23.25-75.19) | -0.190 | 0.849 |
| PCT | 0.055(0.050-0.070) | 0.058(0.050-0.088) | 0.055(0.050-0.070) | -0.520 | 0.603 |
| LYM counts | 1.32(1.00-1.61) | 1.28(0.93-1.80) | 1.33(1.00-1.58) | -0.275 | 0.783 |
| Total T Lymphocyte counts | 740.50(563.50-1073.75) | 629.00(383.00-1073.00) | 743.00(583.00-1078.00) | -0.745 | 0.456 |
| **ECG** | **84** | **22** | **62** |  |  |
| CRP | 8.08(2.63-22.73) | 14.52(2.79-32.71) | 7.72(2.56-20.26) | -0.905 | 0.365 |
| IL-6 | 4.59(1.82-11.80) | 8.70(1.70-16.54) | 4.10(2.13-8.89) | -1.186 | 0.236 |
| ESR | 42.25(23.25-74.50) | 49.30(20.06-77.52) | 42.25(22.83-73.20) | -0.026 | 0.979 |
| PCT | 0.055(0.050-0.070) | 0.064(0.055-0.085) | 0.050(0.050-0.068) | -2.859 | 0.004** |
| LYM counts | 1.32(1.00-1.61) | 1.06(0.81-1.55) | 1.35(1.08-1.73) | -2.040 | 0.041* |
| Total T Lymphocyte counts | 740.50(563.50-1073.75) | 722.00(536.75-1063.00) | 753.00(555.25-1125.00) | -0.621 | 0.535 |

Abbreviations: CRP, C-reactive protein; ESR, erythrocyte sedimentation rate; PCT, procacitonin; IL-6, interleukin 6; cTnI, cardiac troponin I; ECG, electrocardiogram; <sup>a</sup>*P* values indicate differences between severe and nonsevere types of patients. Variables were expressed as median (Q1-Q3).
\**P*<0.05; \*\**P*<0.01

### Multivariate logistic regression analysis

In this study, multivariable logistic regress model was used to identify the independent risk factors for clinical severity of COVID-19 (figure 6). Omnibus Tests of Model Coefficients indicated that the χ^2^ of the multivariate logistic model was 32.595 (p<.050), validating the statistical significance of the model. We tried to include several indicators with statistical differences amongst univariate analyses (except laboratory tests), and the multicolinearity test was carried out according to variance inflation factor. According to the equation fitting degree and the independence of indicators, 6 variables were finally included in multivariable logistic regress model including age, PII, ct value, anorexia, and diabetes. The final results determined that age [OR 2.350; 95% CI (1.206 to 4.580); p =0.012], Ct value [OR 0.158; 95% CI (0.025 to 0.987); p=0.048] and PII [OR 1.912; 95% CI (1.187 to 3.079); p=0.008] were statistically different--independent risk factors for COVID-19(table 7).

**Table 7.**
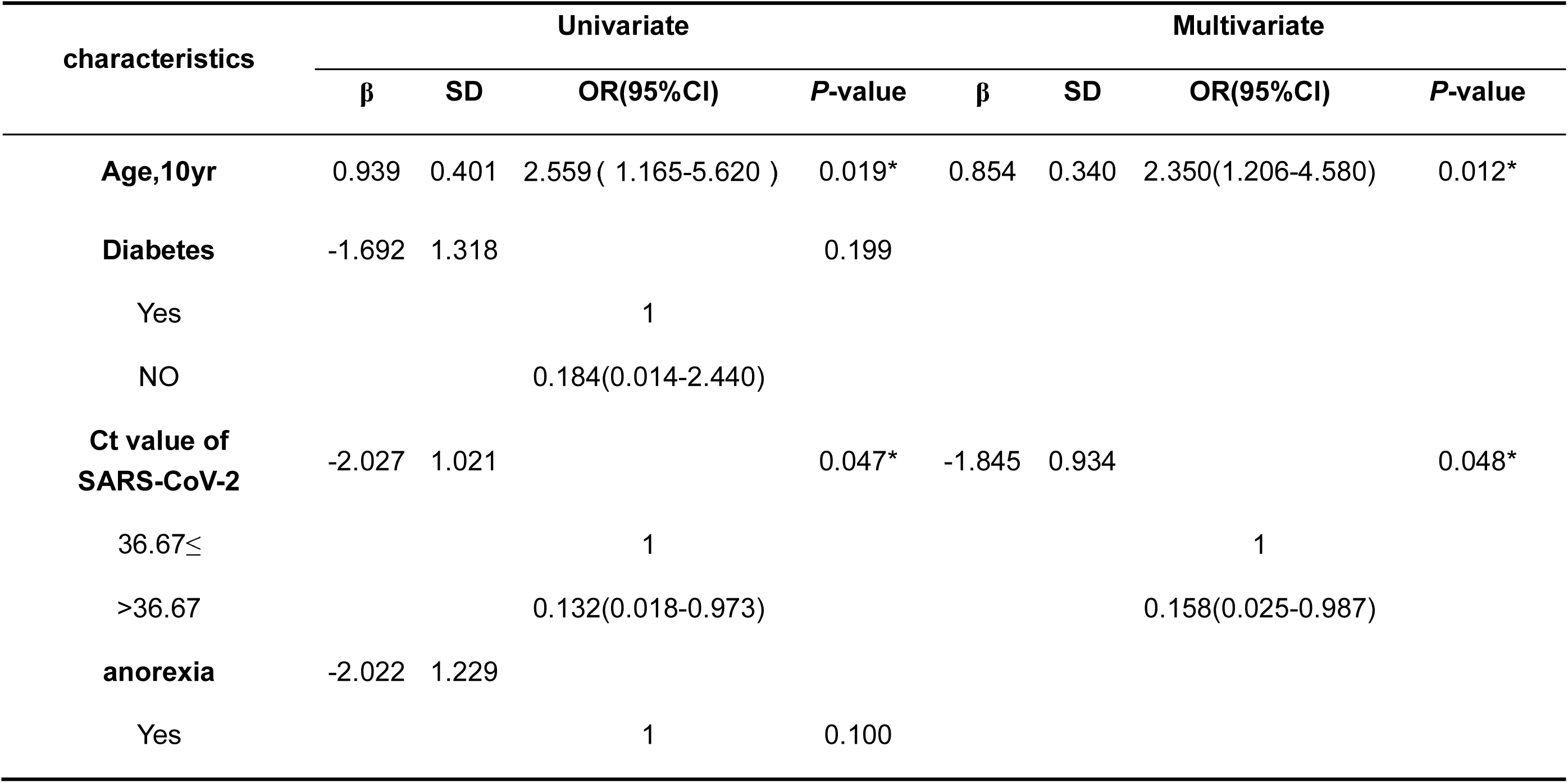
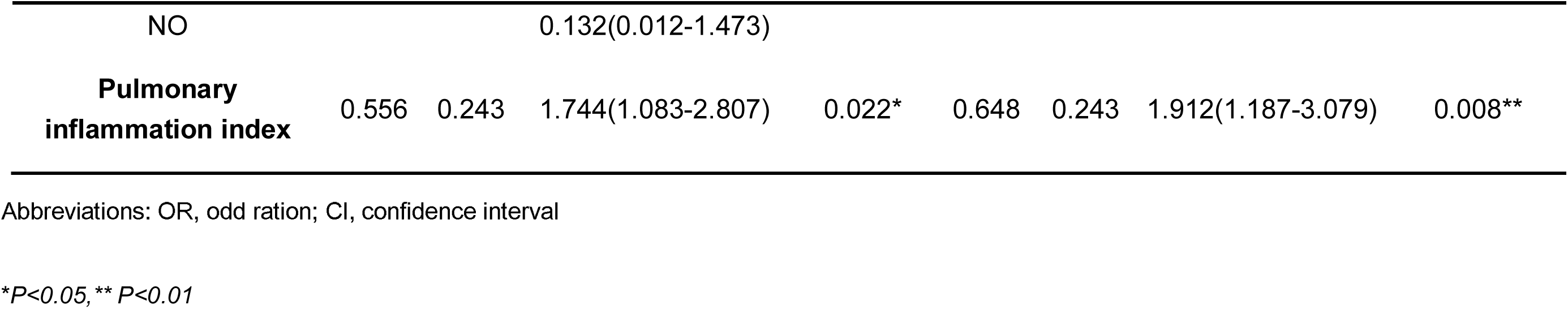
Multivariate Logistic Regression Analysis of Risk Factors.

**Figure 6.**
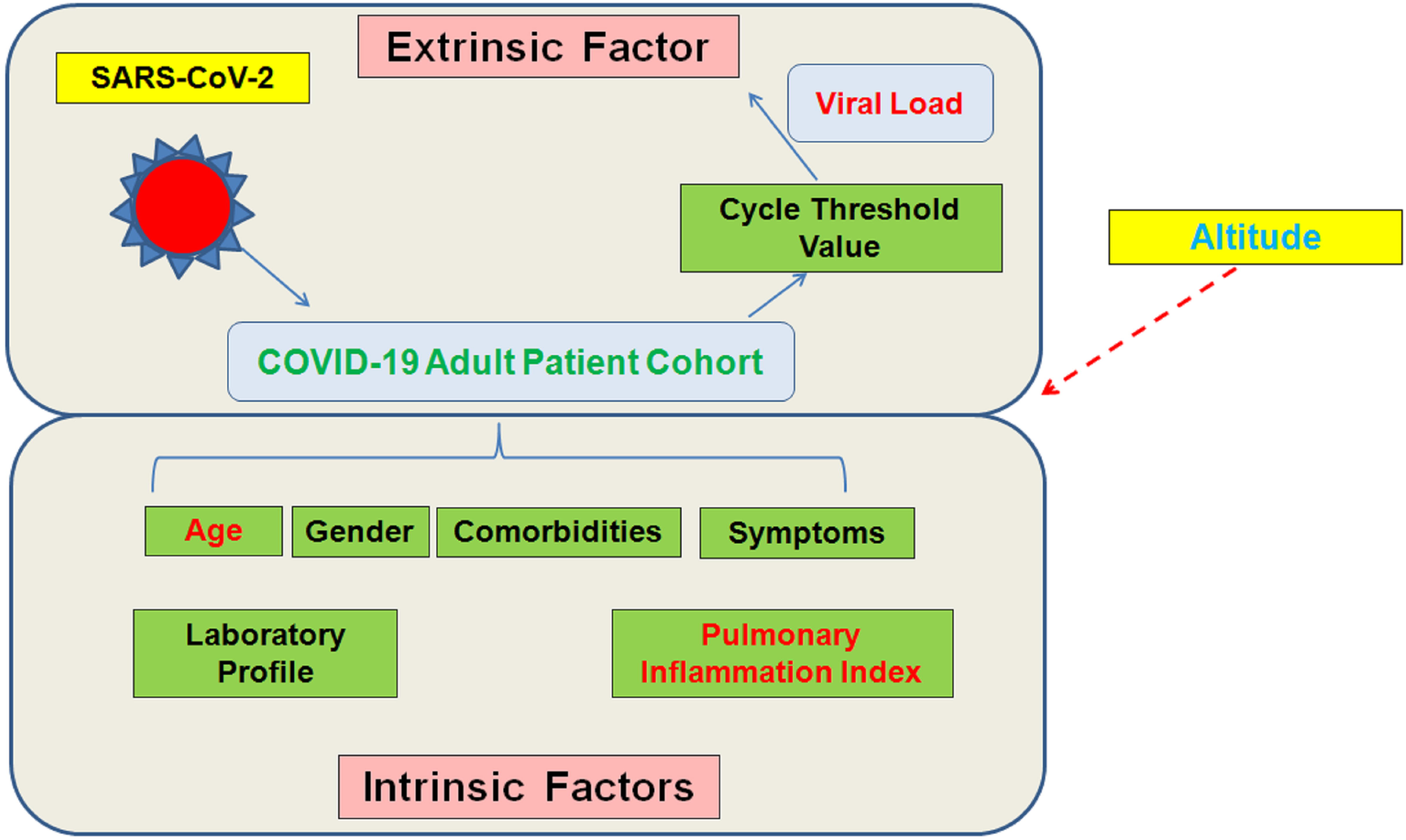
The extrinsic factors and intrinsic factors of SARS-CoV-2.

## Discussion

To our knowledge, this is the first clinical cohort study to analyze the extrinsic (viral load) and intrinsic risk factors of patients with COVID-19. Moreover, SARS-CoV-2 myocarditis was diagnosed clinically with an incidence rate of 4.8%. Previous neglected regional altitude issue was considered for COVID-19 severity classification.

Previous studies mainly focused on the clinical characteristics, imaging findings and treatment measures of COVID-19,^2,3,5-8^ however, few studies pay attention to its risk factors and complications.^14-16^ There were significant differences between severe and nonsevere COVID-19 in age, clinical manifestations, imaging findings, levels of inflammatory cytokines and myocardial enzymes, and distribution of lymphocytes and their subsets, consistent with previous studies.^3,5-8^ Furthermore, multivariable logistic regress analysis identified three key independent risk factors for the severity of COVID-19, i.e., age, PII, and Ct values.

Ct values from the RT-PCR can act as a proxy for viral load.^17,18^ In viral respiratory infectious diseases, viral load has been proved to be closely related to the severity of the diseases.^19-21^ It is still unclear whether the virus load is closed related to the severity of COVID-19 patients, although sporadic case studies have been reported^18,22^. We found that the patients’ viral load was associated with severity of COVID-19 as an independent risk factor.

In addition, all the complications (except for gastrointestinal disorders) of COVID-2019 during hospitalization were found significantly higher in severe than nonsevere patients. Among all patients, a total of 36 patients had a high level of cardiac enzymes or cTnI. After excluding the history of cardiac diseases and combining the ECG and clinical manifestations, 4 patients were clinically diagnosed with SARS-CoV-2 myocarditis according to most updated diagnostic criteria,^12,13^ of which 3 cases occurred in severe group and 1 in nonsevere group. However, the pathological mechanism by which the COVID-19 causes viral myocarditis is still uncertain.

Common pathogens for myocarditis include viruses as coxsackievirus B3 (CVB3),^23^ belonging to the Enterovirus genus in the family of Picornaviridae. CVB3 accounts for over 33% of myocarditis patients.^24^ Kimura et al^25^ reported that IFITs (IFN-induced tetratricopeptides) play a basic role in cardiomyocytes’ antiviral process. IFITs comprise five human sub-family (*IFIT1, IFIT2, IFIT3, IFIT5 and IFIT1B*) members.^26^ Tomko et al^27^identified the coxsackievirus and adenovirus receptor (CAR) in 1997 and confirmed by Bergelson et al^28^ in 2005. CVB3 interacts via the coreceptor decay accelerating factor (DAF) with human cells.^29^ As a type I transmembrane protein, CAR localizes in the tight junctions of epithelial cells and in the intercalated discs between cardiomyocytes^30^. ACE2 has been predicted as receptor for SARS-CoV-2 in case of the similarity with SARS-CoV. The issue remains elusive as for SARS-CoV-2 directly interacting with CAR and damaging myocardiocytes. These issues need to be studied by further evidence.

At present, there is no uniform standard for severity classification of COVID-19. National Health Commission of China (5^th^ edition)^9^ mainly classified the severity classification of COVID-19 according to the respiratory condition of patients, such as respiratory rate (RR) >30 times /min, oxygen saturation<93% at rest, PaO2/FIO2 ratio<300mmHg. The patient’s respiratory rate and blood oxygen saturation are affected not only by the disease itself, but by the altitude of the location. The higher the altitude, the lower the atmospheric pressure and the partial oxygen pressure, and *vice versa*. Pérez et al investigate the prevalence of oxygen desaturation in adults aged ≥40 years as altitude above sea level increases.^31^ They concluded that altitude was closely related to hypoxaemia and might be the most important determinant of low oxygen saturation, inspite of other known factors such as age, diabetes and lung diseases. In subsequent study, Pérez et al^32^ also found that higher altitude of residence was associated with a higher risk of hospitalization and death in patients with influenza-like illness and severe acute respiratory illness. However, neither the National Health Commission of China (5^th^ edition)^9^ nor the American Thoracic Society guidelines^11^considered the influence of the altitude of the pathogen site on the oxygen pressure. In our study, we calculated the correct PaO2/FIO2 ratio by taking altitude into account, which was conducive to accurate classification and subsequent treatments.

There are some limitations in our study. Firstly, the sample size in our study was relatively small, only 84 patients were included. The deadline of the collected data was March 2, 2020, with 16 patients still hospitalized. Secondly, we did not get exact viral load of patients as viral copy numbers due to the urgent outbreak of COVID-19 and limited conditions. We used Ct values as a substitute. Nevertheless, the results of multivariable logistic regress model demonstrated that the Ct value of SARS-CoV-2 is valid and reliable. Thirdly, we diagnosed four cases of viral myocarditis clinically, without evidence of cardiac biopsy as the gold standard of diagnosis. The echocardiography and cardiac MRI were unavailable due to limited protective measures. However, we strictly excluded the interference of the patient’s cardiac history, and made a clinical diagnosis based on the clinical manifestations of myocardial injury, high levels of cardiac enzymes and cTn I, as well as typical ECG changes, which were consistent with the most updated diagnostic criteria of viral myocarditis.^12,13^

## Conclusions

Three key-independent risk factors of COVID-19 were identified, including age, PII, and Ct value. The Ct value is closely correlated with the severity of COVID-19, and may act as a predictor of clinical severity of COVID-19 in the early stage. SARS-CoV-2 myocarditis should be highlighted despite a relatively low incidence rate (4.8%). The oxygen pressure and blood oxygen saturation should not be neglected as closely linked with the altitude of epidemic regions.

## Data Availability

The datasets used to support the current study are available from the authors upon reasonable request.

## Contributors

HQW and KLM conceptualized the study. MKL, CFC, LXK, KQW, LBH, FJL, JBZ, QLL, SLL, and WGT completed the investigation of cases and collected epidemiological data.JL, HQW, LZ, KLM, ZHL, and JZ analyzed the data, with input from KLM, ZHL, JZ, JL, and CFC. HQW, LBH, WGT, QBW, GMQ, and JBZ provided technical assistance and input in content areas, including infection control, epidemiological methods, medical countermeasures, and subject matter expertise. KLM and HQW wrote the initial draft with all authors providing critical feedback and edits to subsequent revisions. All authors approved the final draft of the manuscript. HQW and QBW are the guarantors. The corresponding author attests that all listed authors meet authorship criteria and that no others meeting the criteria have been omitted.

## Funding

No funding.

## Competing interests

None.

## Ethical approval

This study was approved by the Ethics Review Committee of Yongchuan Hospital, Chongqing Medical University (No. 2020KLS-6).

## Patient consent

Obtained.

## Notes

### Competing Interest Statement

The authors have declared no competing interest.

### Funding Statement

No funding was received for the study.

## References

1. Organization WH. Coronavirus disease (COVID-2019) situation reports-34.: World Health Organization; 2020 [Available from: https://www.who.int/docs/default-source/coronaviruse/situation-reports/20200223-sitrep-34-covid-19.pdf?sfvrsn=44ff8fd3_2 accessed 24 Feb 2020.

2. Zhu N, Zhang D, Wang W, et al. A Novel Coronavirus from Patients with Pneumonia in China, 2019. N Engl J Med 2020;382(8):727–33. doi: 10.1056/NEJMoa2001017

3. Wu F, Zhao S, Yu B, et al. A new coronavirus associated with human respiratory disease in China. Nature 2020 doi: 10.1038/s41586-020-2008-3

4. Zhou P, Yang XL, Wang XG, et al. A pneumonia outbreak associated with a new coronavirus of probable bat origin. Nature 2020 doi: 10.1038/s41586-020-2012-7

5. Wu Z, McGoogan JM. Characteristics of and Important Lessons From the Coronavirus Disease 2019 (COVID-19) Outbreak in China: Summary of a Report of 72314 Cases From the Chinese Center for Disease Control and Prevention. JAMA 2020 doi: 10.1001/jama.2020.2648

6. Wang D, Hu B, Hu C, et al. Clinical Characteristics of 138 Hospitalized Patients With 2019 Novel Coronavirus-Infected Pneumonia in Wuhan, China. JAMA 2020 doi: 10.1001/jama.2020.1585

7. Xu X-W, Wu X-X, Jiang X-G, et al. Clinical findings in a group of patients infected with the 2019 novel coronavirus (SARS-Cov-2) outside of Wuhan, China: retrospective case series. BMJ 2020;368:m606. doi: https://doi.org/10.1136/bmj.m606 [published Online First: 19 February 2020]

8. Guan WJ, Ni ZY, Hu Y, et al. Clinical Characteristics of Coronavirus Disease 2019 in China. N Engl J Med 2020 doi: 10.1056/NEJMoa2002032

9. China NHCo. New coronavirus pneumonia prevention and control program (5th edn). 2020 [5th:[Available from: http://www.nhc.gov.cn/yzygj/s7653p/202002/3b09b894ac9b4204a79db5b8912d4440.shtml accessed 4 Feb 2020.

10. Wu J, Wu X, Zeng W, et al. Chest CT Findings in Patients with Corona Virus Disease 2019 and its Relationship with Clinical Features. Invest Radiol 2020 doi: 10.1097/RLI.0000000000000670

11. Metlay JP, Waterer GW, Long AC, et al. Diagnosis and Treatment of Adults with Community-acquired Pneumonia. An Official Clinical Practice Guideline of the American Thoracic Society and Infectious Diseases Society of America. Am J Respir Crit Care Med 2019;200(7):e45–e67. doi: 10.1164/rccm.201908-1581ST

12. Pollack A, Kontorovich AR, Fuster V, et al. Viral myocarditis--diagnosis, treatment options, and current controversies. Nat Rev Cardiol 2015;12(11):670–80. doi: 10.1038/nrcardio.2015.108

13. Olejniczak M, Schwartz M, Webber E, et al. Viral Myocarditis-Incidence, Diagnosis and Management. J Cardiothorac Vasc Anesth 2020 doi: 10.1053/j.jvca.2019.12.052

14. Liu W, Tao ZW, Lei W, et al. Analysis of factors associated with disease outcomes in hospitalized patients with 2019 novel coronavirus disease. Chin Med J (Engl) 2020.

15. Su VYF, Yang YH, Yang KY, et al. The Risk of Death in 2019 Novel Coronavirus Disease (COVID-19) in Hubei Province. Lancet 2020.

16. Zhou F, Yu T, Du R, et al. Clinical course and risk factors for mortality of adult inpatients with COVID-19 in Wuhan, China: a retrospective cohort study. Lancet 2020.

17. Feikin DR, Alraddadi B, Qutub M, et al. Association of Higher MERS-CoV Virus Load with Severe Disease and Death, Saudi Arabia, 2014. Emerg Infect Dis 2015; 21(11): 2029–2035.

18. Zhao W, He L, Xie XZ, Liu J. The Viral Load of 2019 Novel Coronavirus (COVID-19) has the potential to predict the clinical outcomes. Lancet 2020.

19. Lee N, Chan PK, Hui DS, et al. Viral loads and duration of viral shedding in adult patients hospitalized with influenza. J Infect Dis 2009; 200(4): 492–500.

20. Utokaparch S, Marchant D, Gosselink JV, et al. The relationship between respiratory viral loads and diagnosis in children presenting to a pediatric hospital emergency department. Pediatr Infect Dis J 2011; 30(2): e18–23.

21. Jansen RR, Schinkel J, Dek I, et al. Quantitation of respiratory viruses in relation to clinical course in children with acute respiratory tract infections. Pediatr Infect Dis J 2010; 29(1): 82–84.

22. Kim JY, Ko JH, Kim Y, et al. Viral Load Kinetics of SARS-CoV-2 Infection in First Two Patients in Korea. J Korean Med Sci 2020; 35(7): e86.

23. Fung G, Luo H, Qiu Y, Yang D, McManus B. Myocarditis. Circ Res 2016; 118(3): 496–514.

24. Kuhl U, Pauschinger M, Noutsias M, et al. High prevalence of viral genomes and multiple viral infections in the myocardium of adults with “idiopathic” left ventricular dysfunction. Circulation 2005; 111(7): 887–893.

25. Kimura T, Flynn CT, Alirezaei M, Sen GC, Whitton JL. Biphasic and cardiomyocyte-specific IFIT activity protects cardiomyocytes from enteroviral infection. PLoS Pathog 2019; 15(4): e1007674.

26. Fensterl V, Sen GC. The ISG56/IFIT1 gene family. J Interferon Cytokine Res 2011; 31(1): 71–78.

27. Tomko RP, Xu R, Philipson L. HCAR and MCAR: the human and mouse cellular receptors for subgroup C adenoviruses and group B coxsackieviruses. Proc Natl Acad Sci U S A 1997; 94(7): 3352–3356.

28. Bergelson JM, Cunningham JA, Droguett G, et al. Isolation of a common receptor for Coxsackie B viruses and adenoviruses 2 and 5. Science 1997; 275(5304): 1320–1323.

29. Shafren DR, Bates RC, Agrez MV, Herd RL, Burns GF, Barry RD. Coxsackieviruses B1, B3, and B5 use decay accelerating factor as a receptor for cell attachment. J Virol 1995; 69(6): 3873–3877.

30. Cohen CJ, Shieh JT, Pickles RJ, Okegawa T, Hsieh JT, Bergelson JM. The coxsackievirus and adenovirus receptor is a transmembrane component of the tight junction. Proc Natl Acad Sci U S A 2001; 98(26): 15191–15196.

31. Perez-Padilla R, Torre-Bouscoulet L, Muino A, et al. Prevalence of oxygen desaturation and use of oxygen at home in adults at sea level and at moderate altitude. Eur Respir J 2006; 27(3): 594–599.

32. Perez-Padilla R, Garcia-Sancho C, Fernandez R, Franco-Marina F, Lopez-Gatell H, Bojorquez I. The impact of altitude on hospitalization and hospital mortality from pandemic 2009 influenza A (H1N1) virus pneumonia in Mexico. Salud Publica Mex 2013; 55(1): 92–95.

